# Atrophy-related inflammation CSF markers predict clinical progression in genetic frontotemporal dementia

**DOI:** 10.64898/2026.11.10.26367702

**Authors:** Noah L Shapiro, Kamen Tsvetanov, Kei Onn Lai, Peter Simon Jones, Meira Van Schaik, Emily G Todd, Martina Bocchetta, Arabella Bouzigues, Eve Ferry-Bolder, Eva Cullen, Lucy L. Russell, Phoebe H. Foster, Sophie Farley, Rhian Convery, John C. van Swieten, Lize C. Jiskoot, Harro Seelaar, Daniela Galimberti, Rik Vandenberghe, Robert Laforce, Elizabeth Finger, Rose Bruffaerts, Maxime Bertoux, Thibaud Lebouvier, Eino Solje, Johannes Levin, Giuseppe Di Fede, Alexander Thompson, Isabelle Le Ber, Raffaella Lara Migliaccio, Peter Körtvélyessy, Matthias L. Schroeter, Giancarlo Logroscino, Markus Otto, Zeljko Uzelac, Ignacio Illán-Gala, Johanna Krüger, Benedetta Nacmias, Alexander Gerhard, Tobias Langheinrich, Simon Ducharme, Isabel Santana, Maria Carmela Tartaglia, Mario Masellis, Alexandre de Mendonça, Caroline Graff, Fermin Moreno, Matthis Synofzik, Barbara Borroni, Raquel Sanchez-Valle, Genetic FTD Initiative, GENFI, James B Rowe, Jonathan D Rohrer, Maura Malpetti

## Abstract

Neuroinflammation is increasingly recognised as a key contributor to frontotemporal dementia (FTD) pathophysiology, yet how inflammatory processes relate to neurodegeneration and clinical progression remains poorly defined. This study aimed to identify a multivariate cerebrospinal fluid (CSF) inflammatory signature covarying with brain atrophy, and to determine its relevance to clinical and cognitive outcomes in genetic FTD.

We recruited 234 participants (non-carrier controls n=78, presymptomatic carriers n=115, symptomatic carriers n=41) with CSF, MRI, and clinical (CDR ® + NACC FTLD sum of boxes [CDR-SB]) and executive (Wechsler Adult Intelligence Scale-Revised Digit Symbol Substitution Task [DSST] assessments. A regularised canonical correlation analysis (rCCA) was used to derive a shared protein-brain covariance axis. Participants were followed annually for up to 8.6 years (CDR-SB) and 11.5 years (DSST), and linear mixed-effects models were used to estimate individual rates of clinical progression (CDR slope) and executive decline (DSST slope). The predictive value of the inflammation- and MRI-based profiles for cross-sectional CDR-SB, CDR slope, DSST and DSST slope were tested in linear models. Due to floor effects and violations of parametric assumptions, Bias-corrected and accelerated (BCa) confidence intervals (CI) and bootstrap CIs (95%, 5000 replicates) were utilised as the primary inferential criterion in the 2^nd^-level linear models; effects were considered non-significant if the 95% BCa CI crossed zero.

The rCCA identified a robust inflammation-brain axis (r=0.89, p<0.001), driven by CXCL9, FLT3L, CCL3, and frontotemporal-insular atrophy pattern. In symptomatic carriers, the MRI canonical variate was associated with CDR-SB (β=3.80, CI: [0.64, 6.20]) at baseline, whereas the inflammatory protein canonical variate was predictive of CDR-SB slope (β=0.24, CI: [0.02, 0.46]). Whereas both MRI (*β*= −7.27, CI:[−14.20, −1.00]) and protein (*β*= −7.83, CI:[−14.00, −2.16]) canonical variates were linked to DSST cross-sectionally, but not longitudinally. No significant associations were found in pre-symptomatic carriers or non-carrier controls.

A data-driven CSF inflammatory signature, covarying with frontotemporal-insular atrophy, tracks with clinical severity and cognitive impairment in people with genetic FTD, highlighting inflammation as a candidate therapeutic target and progression biomarker in the earliest clinically manifest stages of disease.

## Introduction

Frontotemporal dementia (FTD) encapsulates a heterogeneous group of clinical syndromes linked to neurodegenerative diseases, ranging from pronounced language symptoms (Primary Progressive Aphasia [PPA])^1^ to behavioural changes (behavioural variant FTD [bvFTD])^2^, and motor problems^3^. Approximately 10-20% of people with FTD carry an autosomal pathogenic mutation^3^ that cause the disease, referred to as genetic FTD. The most common mutations include the hexanucleotide (GGGGCC) repeat expansion in Chromosome 9 open reading frame 72 (*C9orf72*)^4^ ^5^, a loss-of-function mutation in progranulin (GRN) causing haploinsufficiency ^6^ ^7^, and missense and splice site mutations in the microtubule-associated protein tau (MAPT)^8^. *C9orf79* and *GRN* mutations are related to TDP43 pathology, while MAPT mutations are linked to tauopathies^9^. Presymptomatic people carrying these genetic mutations show marked changes that can be detected years prior to symptom onset in imaging^10^ ^11^, cognition and behaviour^10^ ^12^ ^13^ ^14^, or proteomic profiles^15^ ^16^.

Neuroinflammation is now recognised as a central and dynamic component of the pathogenesis of FTD^17^, and is associated with faster clinical decline^18^ ^19^. This disease process reflects a highly coordinated cascade of cellular and molecular events involving central nervous system-resident glia (astrocytes and microglia), peripheral immune cells (including monocytes and lymphocytes), and diverse signalling mediators such as cytokines and chemokines^17^ ^20^ ^21^ ^22^ ^23^. Dysregulated cytokine secretion is a key feature of FTD, with converging evidence from blood, CSF, and post-mortem brain tissue^24^ ^25^ ^26^ ^23^. In a retrospective cohort of genetic FTD, Fenoglio et al (2024)^27^ showed that several plasma cytokines, including Tumour Necrosis Factor (TNFα) and interleukins (IL-7, IL-15, IL-16, IL-17A) are significantly upregulated in symptomatic *MAPT* and *GRN* mutation carriers^27^. Complementing these findings, large-scale CSF proteomic studies in genetic FTD have demonstrated that the three most common mutation groups exhibit distinct pathogenic signatures, with immune-related pathways particularly enriched in symptomatic MAPT carriers^28^. In sporadic FTD, multiple studies report upregulation of TNFα, interleukins and chemokines in blood and CSF relative to unaffected controls^29^ ^30^. In sporadic FTD, proinflammatory cytokines in blood have been found to be strongly associated with central inflammation and shorter survival^23^. Taken together, there is substantial evidence for dysregulated immune pathways in FTD, but their relationships with clinical trajectories and neurodegeneration remain incompletely understood. In a large multicentre FTD consortium study of more than 390 participants, Asken and colleagues^31^ quantified IL-6, TNFα and YKL-40 in plasma from presymptomatic and symptomatic mutation carriers and healthy controls, directly linking inflammatory markers to disease progression. Higher TNFα levels were associated with faster clinical decline and temporal lobe atrophy, both cross-sectionally and longitudinally, in MAPT carriers, whereas in C9orf72 carriers, elevated TNFα was related to accelerated cognitive and clinical de-cline without a clear longitudinal atrophy signature. These genotype-specific associations highlight the need to better characterise how inflammatory protein abundance maps onto patterns of neurodegeneration across the FTD spectrum. In line with this, Ullgren and colleagues reported that individual immune-related CSF proteins, such as SERPINA3, are associated with brain atrophy and clinical severity in people with FTD^32^. However, no study has yet captured the multidimensional structure of immune-associated multi-protein profiles and their covariance with atrophy patterns and clinical changes over time, leaving a critical gap in our understanding of immune-brain coupling in FTD. Univariate statistical approaches often cannot capture complex links between modalities, and analysing multidimensional patterns allows us to potentially derive clinically relevant therapeutic target(s) even in presymptomatic stages.

In this study, we aimed to identify a data-driven proteomic profile associated with regional atrophy in genetic FTD, using multivariate statistics of CSF and MRI data from the Genetic FTD Initiative (GENFI) study. We evaluated whether atrophy-related inflammatory signatures are associated with clinical and executive decline, under the hypothesis that greater inflammatory burden would track with faster deterioration. We also tested whether these inflammation-atrophy-clinical relationships are already detectable in the presymptomatic phase by comparing healthy non-carrier controls with presymptomatic and symptomatic mutation carriers.

## Materials and methods

### Participants

From the GENFI (Genetic Frontotemporal dementia Initiative [https://www.genfi.org]) study (data freeze 8)^10^, n=234 participants who underwent CSF sampling and volumetric T1-weighted MRI acquisition at the same visit, as well as standardised GENFI assessments, were included in this study. Specifically, we included n=41 symptomatic carriers (9 with mutation in *MAPT*, 12 in *GRN*, and 20 in *C9orf72*), n=115 pre-symptomatic (defined as the absence of a formal diagnosis) mutation carriers (23 with mutation in *MAPT*, 40 in *GRN*, and 52 in *C9orf72*), and n=78 family members without mutations (non-carrier control group). Participants with another significant medical or psychiatric condition that would interfere with their completion of assessments or impair their safety in the study, or participants who were pregnant or had contraindications to MRI, were excluded.

### Clinical Outcomes

Participants underwent clinical and cognitive assessments at baseline and annually for up to 11.5 years.

To evaluate clinical decline, we utilised the Sum of Boxes score from the CDR+NACC FTLD, as it has been widely used in FTD clinical trials as primary clinical outcome^33^ ^34^. The CDR® +NACC FTLD captures eight domains, which are particularly affected in FTD phenotypes^35^ ^36^ ^35–37^. The CDR® +NACCOFTLD (referred to as CDR-SB onwards) ranges from 0-24, where higher scores indicate more severe symptomology ^38^.

Executive decline was indexed using the Wechsler Adult Intelligence Scale-Revised Digit Symbol Substitution Task (DSST)^39^,ranging from 100 (no executive dysfunction) to 0 (executive dysfunction). For more details on clinical outcomes, see supplementary methods.

### Imaging data acquisition and preprocessing

MRI acquisition has been described elsewhere^40^. Briefly, T1-weighted volumetric (1.1 mm isotropic resolution) scans were acquired on 3 Tesla scanners (Siemens Trio [(n=59], Siemens Skyra [n=32], Siemens Prisma [n=92], Philips Achieva [n=49], GE Discovery MR750 [n=2]) under the GENFI MRI protocol and underwent visual quality controls by the imaging core team. For more details on MRI processing, see supplementary methods.

### CSF sample collection and processing

CSF samples were collected by lumbar puncture into polypropylene tubes and stored at −80 °C at each site. CSF proteins were measured using the proprietary Olink® Target 96 platform. For more details on the processing of the proteomic data, see supplementary methods.

### Statistical analyses

Statistical analyses were performed in R version 4.5.2 and MATLAB (version R2023b).

For descriptive statistics, chi-square and analysis of variance tests compared variables between groups.

#### Canonical Correlation Analysis of the inflammation-atrophy profile

To investigate the inflammation-brain covariance axis, we applied a two-level approach previously deployed in our studies^41^ ^42^ ^43^ ^12^ ^23^. In the first step, we applied a regularised Canonical Correlation Analysis (rCCA) in MATLAB with classical bootstrapping (1000 replicates) to ensure solution stability (supplementary methods for more details on rCCA). In the second step, we imported the canonical variates (CV) and associated loadings from the previous step into R and constructed a linear model with the “lm” function to model MRI-CV using an interaction term between protein-CV and groups (factor with the following levels; controls=0, presymptomatic carriers=1, and symptomatic carriers=2) as the predictor, with age, sex and scanner model as covariates (*MRI-CV∼Protein-CV*Group+Age+Sex+scanner model).* Next, we tested for group differences in the resulting CV variables from the protein and MRI data and ran an Analysis of Covariance (ANCOVA) controlling for age and sex. The ANCOVA was implemented as a linear model using the “lm” function, tested with Type III ANOVA from the car package, and post hoc contrasts were computed using the emmeans package with Bonferroni correction.

#### Second^-^level analyses linking the inflammation-atrophy axis to clinical outcomes

To estimate longitudinal trajectories, we fitted linear mixed-effects models (lme4 “lmer”) to CDR-SB and DSST scores separately, modelling each as a function of years since baseline (MRI/CSF visit) with random intercepts and slopes per participant. Participant-specific slopes (CDR slope, DSST slope), and cross-sectional scores and slopes were then compared across the three clinical groups using one-way ANOVAs (supplementary Figure 2). The predictive value of the protein and MRI latent factors (or CVs) on clinical outcomes was tested in linear models using the CDR-SB, CDR-SB-slope, DSST, and DSST-slope as dependent variables, and CV values*group (factor levels: controls=0, presymptomatic carriers=1, symptomatic carriers=2) as an interaction term. First, we ran separate models with the protein-CV and MRI-CV as single predictors. Then, we tested the combined predictive values of the two variables in a three-way interaction model (protein-CV*MRI-CV*Group). Age, sex, years of education and scanner model were entered as covariates, with controls as the reference group. To directly test whether the association between the canonical variates and outcomes also differed between symptomatic carrier and presymptomatic carrier groups, the group variable was relevelled with presymptomatic carriers as the reference category (factor levels: presymptomatic carrier=0, control=1, symptomatic carriers=2) and the above models were refitted (referred to as “relevelled model”).

#### Inflammation protein signatures independent of atrophy

To isolate the protein-only signal from the atrophy-linked protein profiles (protein-CV), we residualised the 92 proteins. For more details on residualisation analyses and consequent application of PCA’s, see supplementary methods

#### Symptomatic-carrier group-specific analyses

Within the symptomatic carrier group, we also tested associations between clinical outcomes and both the residualised protein component and the protein-CV using non-parametric Spearman’s rank correlations. Given the smaller sample size of the symptomatic carrier subgroup, we additionally computed one-tailed Bayesian Spearman’s correlations (https://osf.io/gny35/), which enable a more robust statistical interpretation by computing Bayes Factors (BF). These correlations were complemented by linear models including covariates.

#### Group stratification by clinical severity

To better capture presymptomatic clinical spectrum effects, we re-stratified groups by CDR-SB score rather than diagnosis alone, following previously established criteria for mutation carriers ^40^ ^10^ ^36^: asymptomatic (CDR-SB=0, n=78), prodromal (CDR-SB=0.5, n=19), and symptomatic (CDR-SB≥1, n=44). Models were re-run substituting disease status with clinical stage (controls=0, asymptomatic=1, prodromal=2, symptomatic=3).

Due to floor effects and violations of parametric assumptions, Bias-corrected and accelerated (BCa)^44^ confidence intervals (CI), bootstrap CIs (95%, 5000 replicates) were utilised as the primary inferential criterion in the 2^nd^ level linear models using the “boot” function from the boot package: effects were considered non-significant if the 95% BCa CI crossed zero. For the interpretation of the Bayesian approach, we set a BF≥3 to evaluate the evidence supporting the alternative hypothesis (a correlation) under the Lee and Wagenmakers criteria^45^.

## Results

### Participants

Participant characteristics are detailed in Table 1. Within the three groups, males were approximately equally distributed 41% (controls), 48% (presymptomatic carriers), and 56% (symptomatic carriers). Symptomatic participants were significantly older than presymptomatic carriers and controls and had fewer years of education (p<0.001). Of the 41 symptomatic carriers, 2.4% had a clinical diagnosis of AD, 4.9 % ALS, 73 % bvFTD, 2.4% CBS, 4.9 % FTD-ALS, and 12 % PPA.

**Table 1.** Demographic and clinical details of participants.

|  | **Healthy Controls (HC, non-carriers)** | **Presymptomatic Carriers (PMC)** | **Symptomatic Carriers (SMC)** | **P-value** |
|---|---|---|---|---|
| **N** | 78 | 115 | 41 |  |
| **Age (Years mean ± SD)** | 45 (12) | 43 (11) | 62 (8) | F (2,201) = 40.91, p < 0.001 |
| **Sex (Female/ Male)** | 46/32 | 60/55 | 18/23 | X<sup>2</sup>(2) = 0.86, p = 0.6 |
| **Education (Years mean ± SD)** | 15 (3) | 15 (3) | 13 (3) | F (2,201) = 8.34, p<0.001 |
| **Genetic Group** |  |  |  |  |
| C9orf72 | - | 52 | 20 |  |
| GRN | - | 40 | 12 |  |
| MAPT | - | 23 | 9 |  |
| **Primary Clinical Diagno-sis** |  |  |  |  |
| AD | - | - | 1 |  |
| ALS | - | - | 2 |  |
| bvFTD | - | - | 30 |  |
| CBS | - | - | 1 |  |
| FTD-ALS | - | - | 2 |  |
| PPA | - | - | 5 |  |

Clinical data were missing for some participants: controls (n=15), presymptomatic carriers (n=7), and symptomatic carriers (n=8) had no CDR-SB score available at the cross-sectional visit. For DSST, we had no available data for presymptomatic carriers (n=1) and symptomatic carriers (n=3). Consequently, those with missing outcome data have been excluded from the 2^nd^-level analysis.

No batch effects were detected (for more details, see supplementary methods).

### CSF inflammatory proteins and brain atrophy correlate in genetic FTD

The rCCA identified a stable pair of positively correlated CVs (r=0.89, p<0.001, Figure 1C). The protein-CV captures upregulated CSF inflammatory signalling (loadings>0.3, Figure 1B), driven by CXCL9, FLT3L, CCL3, CCL2 (also known as MCP-1), LAP-TGF-Beta 1, CCL25, MMP10, OPG, SIRT2, and IL18 markers. The MRI-CV is loaded onto the frontotemporal-insular regions (loadings<-0.6, Figure 1A). For a full overview of the loadings, see supplementary Material Table 1. The positive correlation between the proteomic pattern and regional Brain volumes indicates a strong link between the two modalities.

**Figure 1:**
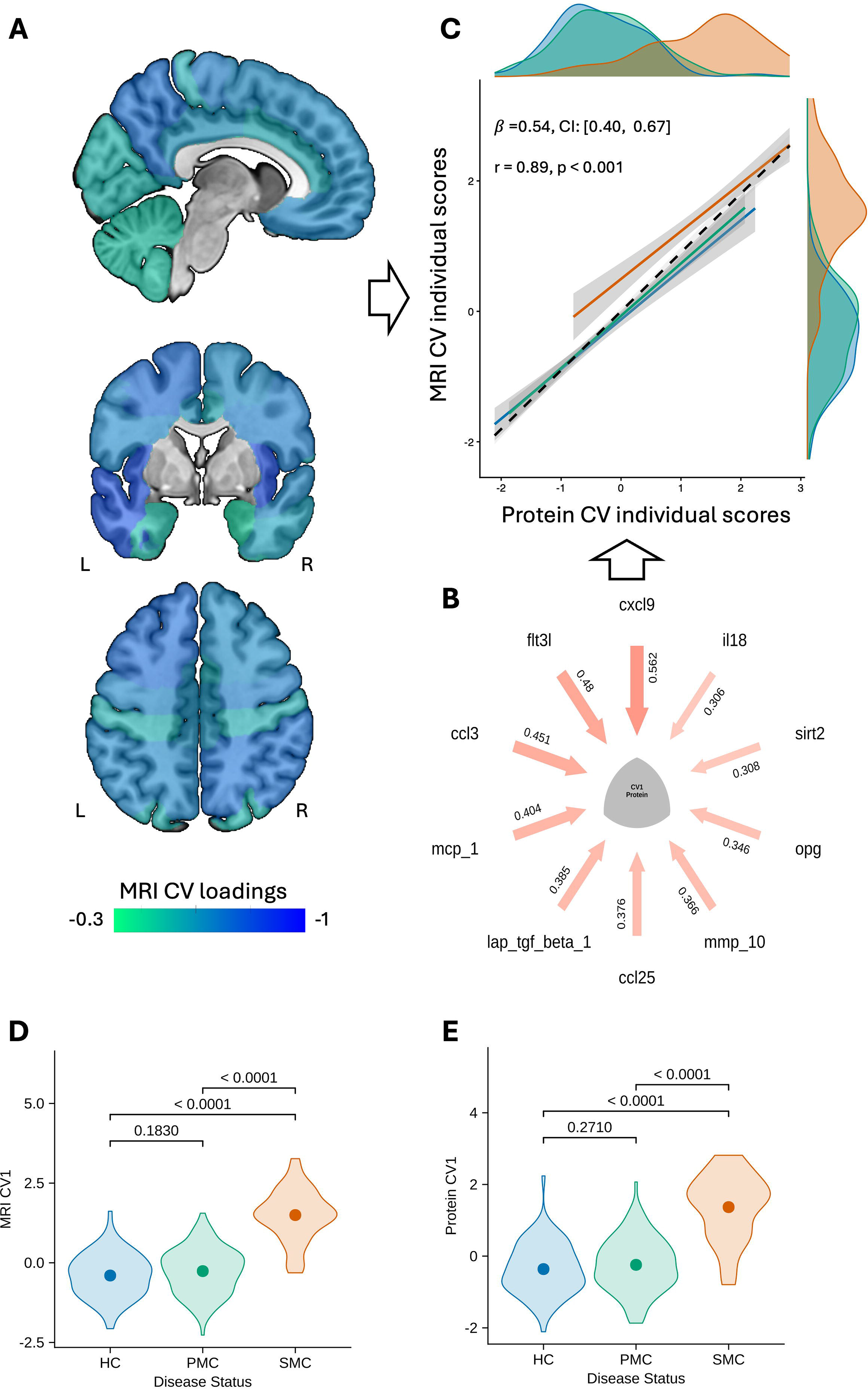
Regularised Canonical Correlation Analysis (rCCA) of CSF inflammatory proteins and MRI regional values. **(A)** Brain plot showing contribution loadings for MRI regional values (representative slice; for a full overview of regional loadings, refer to Supplementary Material 1). Brain regions of interest included the dorsolateral prefrontal (DLPFC), ventromedial prefrontal (VMPFC), orbitofrontal (OF), and motor (MOT) regions. The temporal lobe was subdivided into medial temporal (MT), lateral temporal (LT), temporal pole (TP), and supratemporal areas. The posterior cortical regions were subdivided into sensory (S), medial parietal (MP), lateral parietal (LP), and occipital (OCC). **(B)** Syndromic plot displaying the ten highest-loading CSF proteins contributing to the protein canonical variate (Protein-CV). **(C)** Positive correlation between MRI-derived (y-axis; MRI-CV) and CSF-derived (x-axis; Protein-CV) rCCA participant scores, coloured by disease status (HC = healthy controls [blue], PMC = presymptomatic mutation carriers [mint green], SMC = symptomatic mutation carriers [Vermilion]); the dashed line represents the overall linear model, and marginal plots display density distributions per group. **(D)** Violin plots of MRI-derived CCA participant scores (MRI-CV) by disease status. **(E)** Violin plots of CSF-derived rCCA participant scores (Protein-CV) by disease status. Individual datapoints are not plotted to prevent disclosure of genetic status.

To further disentangle the inflammation-brain axis at each level across the three groups, we fitted a bootstrapped linear model that included an interaction term with group (MRI-CV∼protein-CV*group) including covariates. Protein-CV was significantly associated with MRI-CV (*β*=0.54, CI:[0.40,0.67]), while the interaction terms (protein-CV*group) were not statistically significant for presymptomatic carriers (*β*=0.05, CI:[− 0.09,0.19]) nor symptomatic carriers (*β*=0.09, CI:[−0.09,0.28]). Age was positively associated with MRI-CV scores (*β*=0.01, CI:[0.008,0.02]).

The ANCOVA revealed protein-CV scores differed between groups (F(2,229)=41.57, p<0.001) (Figure 1E). Post hoc comparisons indicated that the symptomatic carriers had significantly higher Protein-CV scores than controls (p<0.001) and presymptomatic carriers (p<0.001). Presymptomatic carriers and controls did not differ (p>0.05). Age and sex were significant covariates (p<0.01). The ANCOVA on the MRI-CV scores identified a similar pattern between groups (F(2,229)=22.92, p<0.001) (Figure 1D), showing that MRI-CV scores in symptomatic carriers were significantly higher than in controls (p<0.001) and presymptomatic carriers (p<0.001), but presymptomatic carriers and controls groups did not differ (p>0.05), with age and sex as significant covariates (p<0.01). Taken together, higher protein-CV scores reflect a pattern of elevated inflammatory signalling related to regional brain atrophy, whereas higher MRI-CV scores reflect greater atrophy across the regions that covary most strongly with the inflammatory profile.

Symptomatic carriers were characterised by significantly higher CDR-SB scores than presymptomatic carriers or controls (p<0.001), while the latter groups did not differ (p=0.79). CDR-SB slopes follow the same pattern. Symptomatic carriers showed significantly lower DSST scores than presymptomatic carriers or controls (p<0.001), but there was no difference between presymptomatic carriers and controls (p=0.68). For DSST slopes, there were no significant differences among the groups.

### CSF inflammatory proteins are associated with clinical progression over time but not with cross-sectional clinical outcomes

The omnibus interaction term between protein-CV scores and disease group in the parametric model on CDR-SB scores was statistically significant (F(2,192)=5.81, p=0.003). However, in the bootstrapped model, the interaction terms were not associated with CDR-SB scores (Ref group: controls (*β*=0.11, CI:[−0.53,0.74]); presymptomatic carriers (*β*=-0.07, CI:[−0.56,0.37]); symptomatic carriers (*β*=1.84, CI:[−0.53,3.71], Figure 2A-right). The relevelled model showed that the association between protein-CV and CDR-SB scores did not differ between symptomatic and pre-symptomatic carriers (*β*=1.96, CI:[−0.5,3.8]). Refitting the bootstrapped linear model using data from symptomatic mutation carriers indicated that protein-CV scores were positively associated with CDR-SB scores (β=1.37, CI:[1.09,3.09], Rho=0.25). However, the BF=1.07 provides only anecdotal evidence for this correlation (Figure 2E-left). In contrast, the omnibus interaction term between MRI-CV scores and disease group in the parametric model was statistically significant (F(2,192)=18.31, p<0.001), concordant with the bootstrapped model, where the interaction terms were associated with higher CDR-SB scores within the symptomatic carriers (*β*=3.80, CI:[0.64,6.20], Figure 2A-left) but not in presymptomatic carriers (*β*=-0.10, CI:[−0.57,0.28]) nor in controls (*β*=-0.007, CI:[−0.66, 0.59]). The relevelled model indicated that the association between MRI-CV and CDR-SB scores differed between presymptomatic carriers and symptomatic carriers (*β*=4.03, CI [0.66,6.49]) on CDR-SB scores. The omnibus three-way interaction term in the parametric model was not significantly (F(2,189)=0.26, p=0.9) associated with CDR-SB scores in concordance with the bootstrapped model (Ref group: controls (*β*=-0.08, CI:[−0.42, 0.27]); pre-symptomatic carriers (*β*=0.08, CI:[−0.38,0.44]); symptomatic carriers (*β*=-0.11, CI:[−3.13, 2.97]).

**Figure 2:**
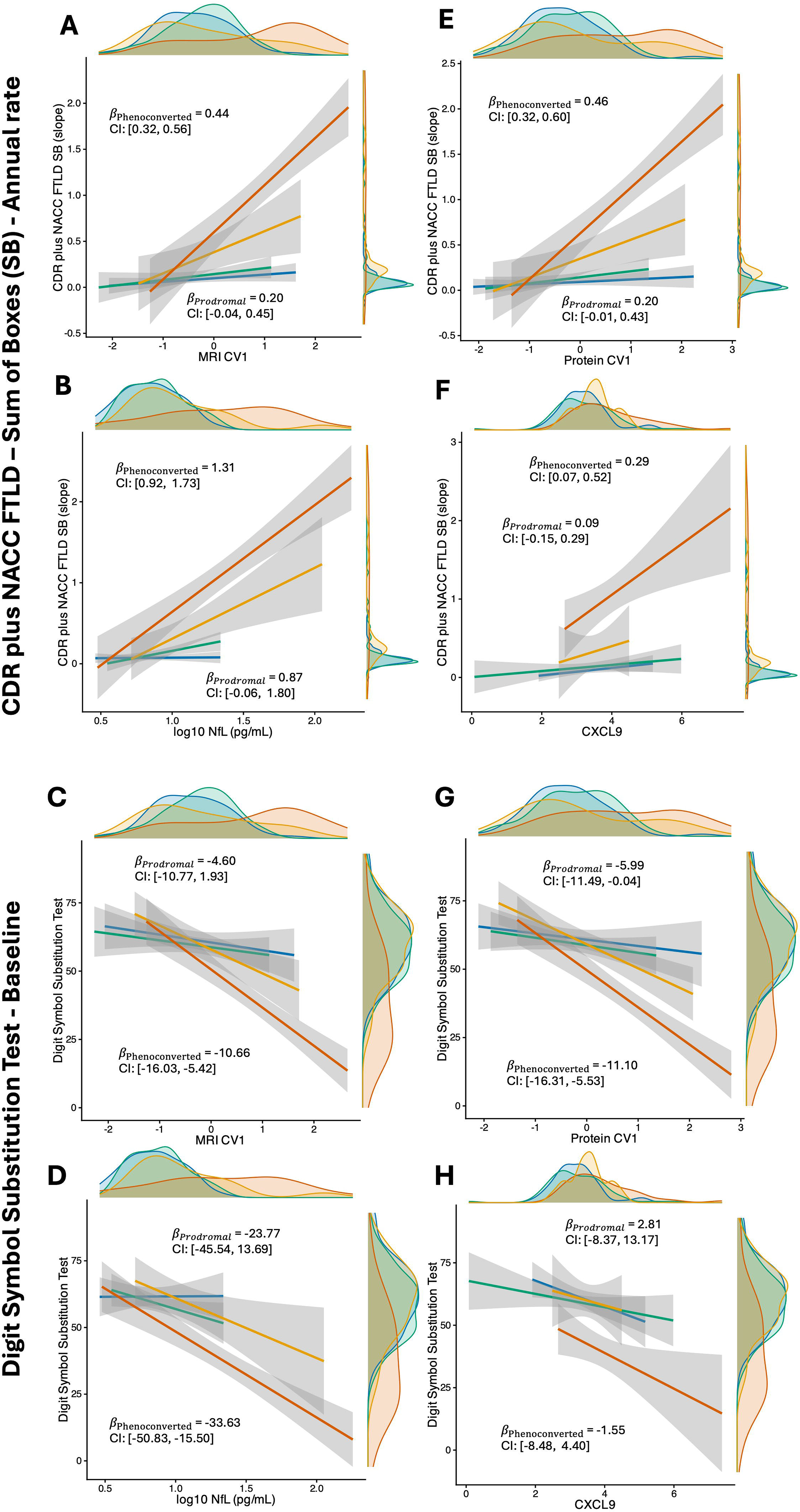
CSF inflammatory protein and MRI canonical variates by clinical and cognitive progression. **(A, E)** Cross-sectional associations with clinical severity (CDR plus NACC FTLD-SB): (A) Primary analysis mapping baseline clinical severity against (left) MRI-CV and (right) Protein-CV scores. (E) Sensitivity validation comparing (left) Protein-CV scores against (right) residualised protein PC1 scores. **(B, F)** Longitudinal analyses of clinical progression (CDR plus NACC FTLD-SB slope): (B) Primary analysis mapping participant trajectories against (left) MRI-CV and (right) Protein-CV scores. **(F)** Sensitivity validation comparing (left) Protein-CV scores against (right) residualised protein PC1 scores. **(C, G)** Cross-sectional associations with executive functioning (Digit Symbol Substitution Test): (C) Primary analysis mapping baseline executive function against (left) MRI-CV and (right) Protein-CV scores. (G) Sensitivity validation comparing (left) Protein-CV scores against (right) residualised protein PC1 scores. **(D, H)** Longitudinal analyses of executive deterioration (DSST slope): (D) Primary analysis mapping longitudinal rates of cognitive change against (left) MRI-CV and (right) Protein-CV scores. (H) Sensitivity validation comparing (left) Protein-CV scores against (right) residualised protein PC1 scores. Individual datapoints are not plotted to prevent disclosure of genetic status. For all panels, linear regressions map associations with regression coefficients and 95% bias-corrected and accelerated confidence intervals (95% BCa CI) reported only for symptomatic mutation carriers. Marginal plots show density distributions by disease status (HC = healthy controls [blue], PMC = presymptomatic mutation carriers [mint green], SMC = symptomatic mutation carriers [vermilion]).

The omnibus interaction term between protein-CV scores and disease group in the parametric model with CDR-SB-slopes was statistically significant (F(2,192)=5.59, p=0.004), Ref group: controls), concordant with the bootstrapped model, where the interaction terms were associated with CDR-SB-slopes in symptomatic carriers (*β*=0.24, CI:[0.02,0.46], Figure 2B-right), but not for presymptomatic carriers (*β*=0.03, CI:[−0.01, 0.14]), or controls (*β*= 0.03, CI:[−0.04, 0.09]). The relevelled model showed that the association between protein-CV and CDR-SB-slopes was not different between symptomatic and presymptomatic carriers (β=0.18, CI:[−0.04, 0.39]. Refitting the bootstrapped linear model using only data from symptomatic mutation carriers indicated that protein-CV scores were positively associated with CDR-SB-slopes (β=0.23, CI:[0.20,0.45], Rho=0.34, BF=3.28, Figure 2F-left). In contrast, the omnibus interaction term between MRI-CV scores and disease group onto CDR-SB-slopes in the parametric model was statistically significant (F(2,192)=3.87, p=0.02, Ref group: controls). However, in the bootstrapped model, the interaction terms were not associated with CDR-SB-slopes (Ref group: controls (*β*=0.03, CI:[−0.05,0.09]; presymptomatic (*β*=0.03, CI:[−0.03,0.11]; and symptomatic carriers (*β*= 0.24, CI:[−0.03,0.53], Figure 2B-left). The relevelled model showed that the association between MRI-CV and CDR-SB-slopes did not differ between symptomatic and presymptomatic carriers (β=0.21, CI:[−0.07,0.50]). The omnibus three-way interaction term in the parametric model was not associated (F(2,189)=0.13, p=0.86) with CDR-SB-slopes in concordance with the bootstrapped model (Ref group: controls *β*=-0.005, CI:[−0.05,0.05]; presymptomatic carriers *β*= 0.007, CI:[−0.07,0.09]; symptomatic carriers (*β*=-0.032, CI:[−0.35,0.20]).

### Both CSF inflammatory proteins and fronto-insular atrophy are associated with worse executive function cross-sectionally

The omnibus interaction term between protein-CV scores and disease group in the parametric model was statistically significant (F(2,217)=4.19, p=0.02) with DSST scores. This is concordant with the bootstrapped model where the interaction term in symptomatic carriers is linked with DSST scores (*β*=-7.83, CI:[−14.00,-2.16], Figure 2C-right) but not for the presymptomatic carriers (*β*=-1.80, CI:[−6.07,2.33]) nor controls (*β*=1.51, CI:[−2.70,5.63]). The relevelled model showed no association between protein-CV and DSST scores between symptomatic and presymptomatic carriers (β=-5.2, CI:[−10.97,0.10]). Refitting the bootstrapped linear model using only data from symptomatic mutation carriers indicated that protein-CV scores were associated with worse DSST scores (β=-6.09, CI:[−11.56,-2.19], Rho=-0.40, BF=44.4, Figure 2G-left). Furthermore, the omnibus interaction term between MRI-CV scores and disease group in the parametric model was statistically (F(2,217)=3.12, p=0.04) as-sociated with DSST scores. This is concordant with the bootstrapped model showing that MRI-CV scores were statistically associated with DSST scores in symptomatic carriers (*β*=-7.27, CI:[−14.20,-1.00], Figure 2C-left) but not for presymptomatic carriers (*β*=-1.07, CI:[−5.40,3.28]) nor controls (*β*=0.54, CI:[−4.09,4.78]). The relevelled model showed that the association between MRI-CV and DSST was not different be-tween symptomatic and presymptomatic carriers (β=-5.20, CI:[−11.63,0.49]). The omnibus three-way interaction term in the parametric model was not (F(2,215)=2.03, p=0.13) associated with DSST scores, which is in concordance with the boot-strapped model showing no association with DSST scores (Ref group: controls *β*=-3.73, CI: [−10.32,5.02]; presymptomatic carriers *β*=2.33, CI:[−1.90,7.56]; symptomatic carriers *β*=5.68, CI:[−0.37,12.38]).

Contrary, the omnibus interaction term between protein-CV scores and disease group in the parametric model was not (F(2,217)=0.56, p=0.57) associated with DSST-slope, which is in concordance with the bootstrapped model, where the inter-action terms were not associated with DSST slopes (Ref group: controls (*β*=-0.16, CI:[−0.41,0.13]; presymptomatic carriers (*β*=-0.11, CI:[−0.54,0.15]); symptomatic carriers (*β*=0.07, CI:[−0.29,0.43], Figure 2D-right). The relevelled model indicated that the association between protein-CV and DSST-slopes was not different between symptomatic carriers and presymptomatic carriers (β=0.18, CI:[−0.14,0.57]). Refitting the bootstrapped linear model using only data from symptomatic mutation carriers indicated that protein-CV scores were not associated with DSST-slope (β=-0.16, CI:[−0.45,0.14], Rho=-0.12, BF=0.74, Figure 2H-left). The omnibus interaction term be-tween MRI-CV scores and disease group in the parametric model was not (F(2,221)=2.06, p=0.12) associated with DSST-slopes. In concordance with the bootstrapped model showing that the interactions were not associated with DSST-slopes (Ref group: controls *β*=-0.15, CI:[−0.42,0.13]; presymptomatic carriers *β*=-0.21, CI:[−0.69,0.11]; symptomatic carriers *β*=0.20, CI:[−0.19,0.57], Figure 2D-left). The relevelled model showed that the association between MRI-CV and DSST slopes was statistically different between symptomatic and presymptomatic carriers (β=0.41, CI:[0.06,0.84]). The omnibus three-way interaction term in the parametric model was not (F(2,215)=1.04, p=0.35) associated with DSST-slope (Ref group: controls *β*=0.16, CI:[−0.14,0.38]; presymptomatic carriers *β*=-0.26, CI:[−0.70,0.10]; symptomatic carriers *β*=-0.09, CI:[−0.50,0.31]).

### The atrophy-related CSF inflammation pattern is clinically relevant

To decouple the protein-CV from the MRI pattern within the symptomatic group, we deployed residualisation. The PCA of the residualised protein values identified one principal component (PC) that met the stipulated criteria, explaining 34.4% of the variance. PC1 was positively loaded across the residualised proteins, with the highest proteins largely distinct from protein-CV (supplementary material Figure 3). PC1 scores were orthogonal to protein-CV scores (r=-0.23, CI:[−0.50,0.08]), confirming that residualisation effectively decoupled the signals.

Next, we linked PC1 scores to the same clinical outcomes as the protein-CV scores, but found no statistically significant association with any cross-sectional (CDR-SB: *β*=-0.22, CI:[−0.57,0.15], rho=-0.28, BF=0.97 Figure 2E-right], DSST: *β*=0.73, CI:[− 0.07,1.83], rho=0.25, BF=0.89 [Figure 2G-right]) or longitudinal outcomes (CDR-SB-slopes: *β*=0.004, CI:[−0.03,0.05]), rho=-0.01, BF=0.24) [Figure 2F-right], DSST-slopes: *β*=0.006, CI:[−0.03,0.05]), rho=-0.009, BF=0.17 [Figure 2H-right]). The double dissociation indicates that the atrophy-related inflammation CSF pattern carries more clinical value than the protein pattern on its own. Furthermore, the supplementary analysis showed no statistically significant association between protein-CV and any of the outcomes across the presymptomatic carriers and controls groups (supplementary material table 2).

PCA applied directly to proteomic values in controls and presymptomatic carriers identified one PC that met the abovementioned criteria (explained 37.4% variance), whose scores did not differ significantly between groups (p=0.61), indicating that the proteomic profile of presymptomatic carriers is not distinguishable from that of controls (supplementary material Figure 4B). Furthermore, the loading structures from two PCAs (residualised within symptomatic carriers and unresidualised in presymptomatic carriers /controls) were statistically significantly correlated (r=0.90, p<0.001) (supplementary material Figure 4A), indicating that the residualised PCA within symptomatic carriers reflects a clinical-group-independent covariation structure shared across the remaining cohort, which is distinct from the clinically relevant signal captured in the rCCA.

### Carriers in the prodromal window showed a slight elevation of inflammatory proteins and neurodegeneration markers

The re-stratified groups based on CDR scores reveal that phenoconverted participants had statistically significantly lower DSST scores (p<0.001) compared with those in the prodromal, asymptomatic, and control groups (Figure 3C). In contrast, CDR-SB-slopes was significantly higher in the prodromal stages (p<0.03, Figure 3F). For the protein-CV scores, only phenoconverted individuals had significantly higher scores than the other groups (p<0.001, Figure 3D). A similar pattern emerges for MRI-CV scores, with significantly higher scores in phenoconverted individuals than in the remaining groups (p<0.001, Figure 3A). Interestingly, there is a stepwise non-significant increase in both protein-CV and MRI-CV scores, indicating prodromal changes in the atrophy-related CSF inflammation pattern. We also examined neuro-filament light chain (NfL) abundance, a proxy marker for neurodegeneration (Figure 3B), and CXCL9 levels (Figure 3E) across the re-stratified groups (supplementary results 1.1). Stratification by genetic groups across the clinical continuum is shown in the supplementary material Figure 5.

**Figure 3:**
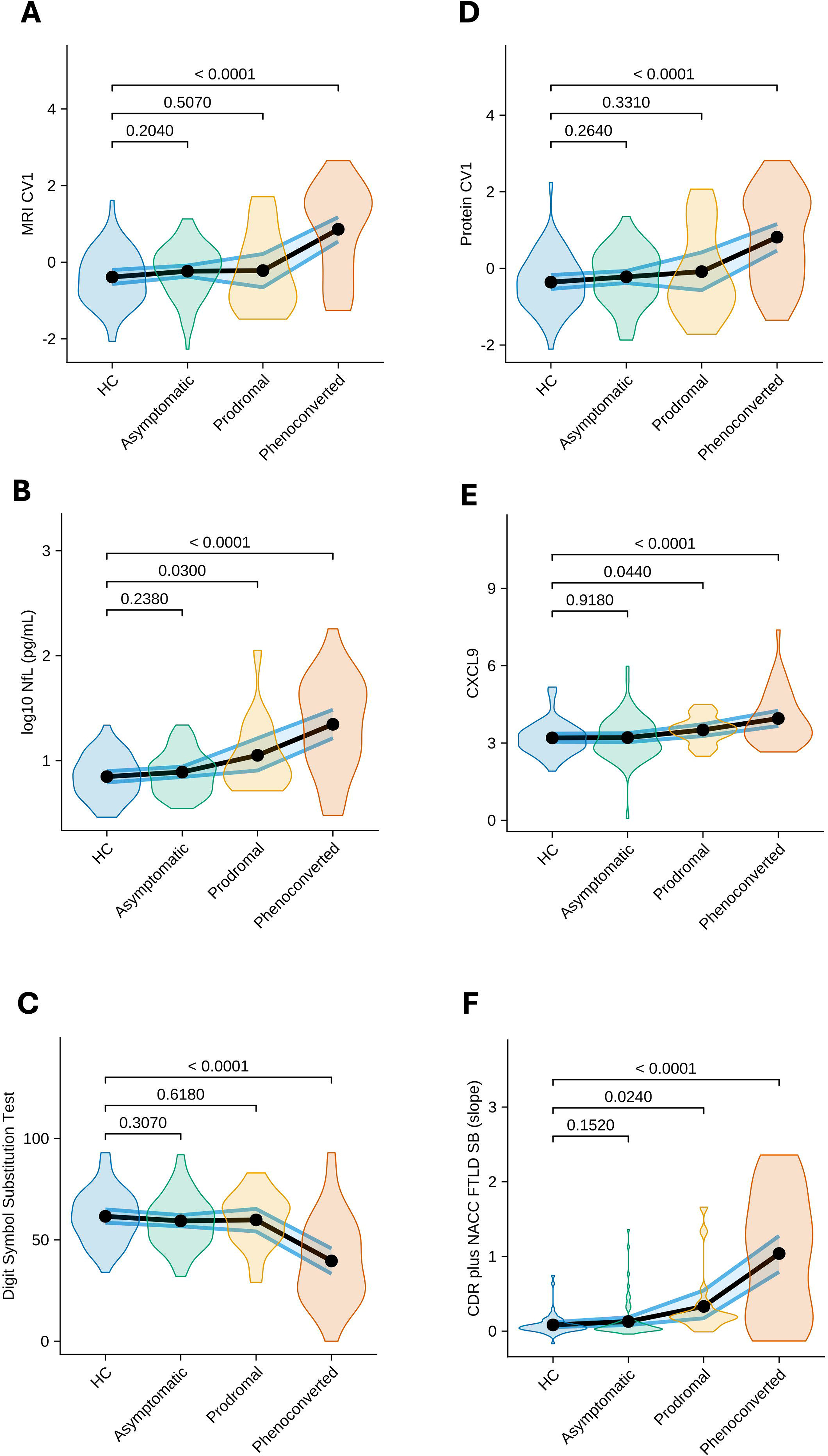
Cognitive, clinical, imaging, and fluid biomarker profiles across the clinical continuum stratified by CDR-SB scores. Participant groups were re-stratified by baseline CDR-SB scores to provide greater sensitivity to the clinical spectrum (Asymptomatic: CDR-SB = 0; Prodromal: CDR-SB = 0.5; Phenoconverted: CDR-SB ≥ 1) alongside healthy controls (HC). Individual datapoints are not plotted to prevent disclosure of genetic status. Violin plots represent data density distributions across the clinical continuum for **(A)** MRI-CV scores, **(B)** log10-transformed Neuro-filament light chain (log10 NfL) levels, **(C)** longitudinal clinical progression captured by the CDR plus NACC FTLD-SB slope (CDR - slope), **(D)** protein-CV scores, **(E)** abundance levels of CXCL9 (the highest loaded protein derived from the Regularised Canonical Correlation Analysis, **(F)** executive function indexed by the Digit Symbol Substitution Test. Black line: tracks group-wise means; Blue lines: 95% CI around the mean.

The omnibus interaction term between protein-CV scores was statistically significant (F(3,190)=19.03, p<0.001), associated with CDR-SB-slopes, which is in concordance with the bootstrapped model, where the interaction terms were significantly associated with CDR-SB-slopes (β=0.46, CI:[0.32,0.60] Figure 4E), whereas the remaining groups were not (controls β=-0.01, CI:[−0.09,0.08], Asymptomatic β=0.06, CI:[−0.02,0.18], Prodromal β=0.20, CI:[−0.01,0.43]). Results on the 2^nd^-level analysis of CXCL9 (Figure 4F; supplementary results 1.2) and NfL levels on CDR slope (Figure 4B; supplementary material results 1.3) are presented in the supplementary material

**Figure 4:**
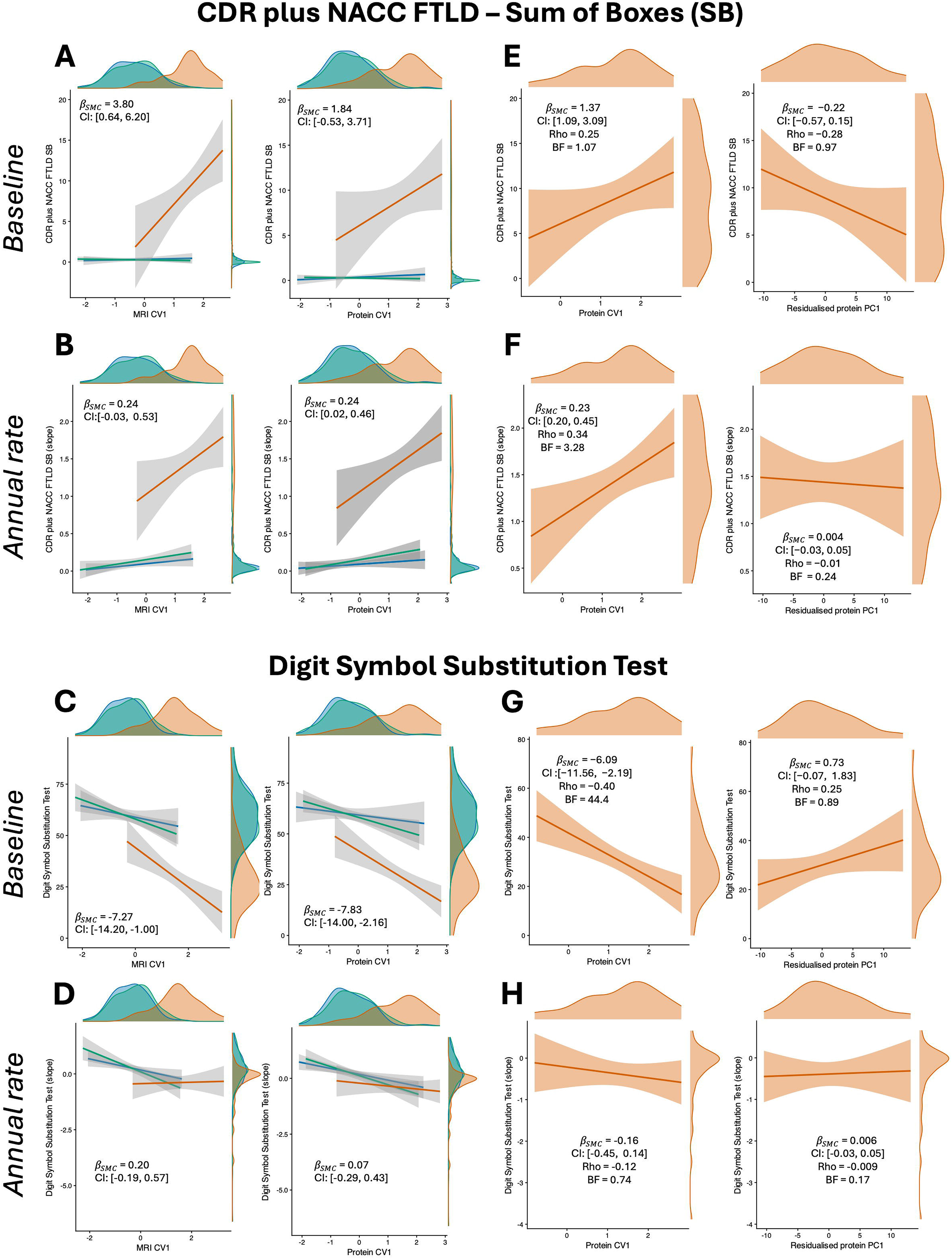
Associations of canonical variates and fluid biomarkers with clinical outcomes across clinical stages. **(A)** Clinical progression (CDR plus NACC FTLD-SB slope) mapped against baseline MRI-CV scores. **(B)** Clinical progression mapped against baseline log10 NfL levels. **(C)** Executive dysfunction (Digit Symbol Substitution Test) mapped against MRI-CV scores. **(D)** Executive dysfunction mapped against log10 NfL levels. **(E)** Clinical progression mapped against baseline Protein-CV scores. **(F)** Clinical progression mapped against baseline CXCL9. **(G)** Executive dysfunction mapped against Protein-CV scores. **(H)** Executive dysfunction mapped against CXCL9. Individual datapoints are not plotted to prevent disclosure of genetic status. All panels display linear regression lines with shaded 95% confidence intervals and marginal density plots displaying distributions by clinical stage (HC = healthy controls [blue], Asymptomatic [bluish-green], Prodromal [orange], Phenoconverted [Vermilion]). Regression coefficients and 95% bias-corrected and accelerated confidence intervals (95% BCa CI) are only reported for the re-stratified prodromal and phenoconverted groups.

The omnibus interaction term between MRI-CV scores was statistically significant (F(3,190)=16.90, p<0.001) associated with CDR-SB-slopes, which is in concordance with the bootstrapped model, where the interaction terms were associated in Pheno-converted participants with worse clinical progression (β=0.44, CI:[0.32,0.56], Figure 4A), whereas the remaining groups were not (controls β=0.02, CI:[−0.07,0.12], Asymptomatic β=0.04, CI:[−0.04,0.15], Prodromal β=0.20, CI:[−0.04,0.45]).

The omnibus interaction term between protein-CV scores and DSST scores were statistically significant (F(3,186)=7.70, p<0.001), which is in concordance with the bootstrapped model, where the interaction terms were significant in phenoconverted and prodromal participants with worse DSST scores (phenoconverted: *β*=-11.10, CI:[−16.31,-5.53], prodromal: β=-5.99, CI:[−11.49,-0.04]). Figure 4G), whereas not for the remaining groups (controls: β=1.65, CI:[−2.85,5.96], Asymptomatic: β=-1.71, CI: [−6.84,3.15], The omnibus interaction term between MRI-CV in the parametric model was statistically significant (F(3,186)=7.42, p<0.001), associated with DSST scores, which is in concordance with the bootstrapped model, where the interaction terms were statistically significant in Phenoconverted participants (β=-10.66, CI:[−16.03,-5.42], Figure 4C), whereas the remaining groups did not (controls: β=1.13, CI:[− 3.79,5.78], Asymptomatic: β=-0.58, CI:[−5.87,4.78], Prodromal: β=-4.60, CI:[− 10.77,1.93]). Results on the 2^nd^-level analysis of CXCL9 (Figure 4H, supplementary results 1.4) and NfL levels on DSST scores (Figure 4D; supplementary results 1.5) are presented in the supplementary material.

## Discussion

Our study integrates multiplex CSF proteomics and MRI to investigate the relationship between inflammatory and atrophy profiles, and their clinical relevance and predictive value in genetic FTD. Our data-driven approach identified a robust inflammation-atrophy covariance axis that predicts clinical decline over time in people with symptomatic FTD. Specifically, the atrophy-related CSF inflammation pattern predicted worse CDR-SB scores over time in symptomatic FTD gene carriers. Inflammatory markers and fronto-insular atrophy measures differentiated symptomatic from presymptomatic and control participants, but could not separate presymptomatic from control participants. To further elucidate protein-brain covariance across the presymptomatic continuum, we re-stratified participants by CDR-SB scores, which more sensitively capture granular clinical changes than the simple absence of a formal clinical diagnosis. When deploying this approach, we found that inflammatory protein and MRI covarying scores trended towards stepwise divergence between the prodromal and asymptomatic stages. Examining single protein levels of both CXCL9 (the highest loaded protein derived from the rCCA) and NfL, we detected statistically discernible increases in prodromal stages. For the 2^nd^-level analyses, both group stratification strategies yielded convergent interpretations. However, within the pro-dromal FTD cohort, associations with executive dysfunction and clinical progression did not reach statistical significance.

Applying a CCA across proteomics and MRI data, we identified an atrophy-related inflammatory profile that was driven by chemokines (CXCL9, CCL3, CCL2 or MCP-1 and CCL25) that are involved in the recruitment of T-cells and Natural Killer cells^46^, monocytes and macrophages^47^, as well as FLT3L, which promotes the development of dendritic, B, and natural killer cells, and MMP10, which is involved in tissue re-modelling^48^ ^49^. The CCA also identified an atrophy pattern linked with inflammation, which encompassed frontal and temporal regions and the insula, consistent with their central and early involvement in FTD^50^ ^51^ ^10^. Notably, interpretation of rCCA-derived canonical variates differs from other dimensionality reduction techniques such as PCA: rCCA assigns weights to maximise correlation between modalities, rather than reducing dimensionality within each modality independently. In this context, higher CSF protein-CV scores indicate increased inflammatory signalling associated with regional brain atrophy, while higher MRI-CV scores reflect more pronounced atrophy in regions that most strongly covary with the inflammatory profile. The primary advantage of rCCA is that it treats inflammation and neurodegeneration as an integrated system, capturing complex multimodal crosstalk within a unified statistical framework.

The atrophy-related inflammation CSF pattern, led by CXCL9, identified by our data-driven analysis was elevated in symptomatic patients. Previous studies focusing on single inflammation markers did not find significant increase in CXCL9 levels in blood and CSF of patients with PPA^52^ or bvFTD^29^. Mechanistically, CXCL9 is an IFN-γ inducible chemokine that signals through the CXCR3 receptor^53^ ^54^, a pathway implicated in T-cell recruitment to the CNS and to sites of inflammation^55^, that potentially contributes to neurodegeneration. On the other hand, multiple studies have shown increased levels of MCP-1 in CSF of people with FTD compared with controls^56^ ^21^. MCP1 (also known as CCL2) is implicated in the pathogenesis of a range of human diseases^57^. MCP1 plays an important role in the inflammatory response, acting as a modulator of leukocytes, and its levels have been linked to AD disease severity^58^ ^59^. MCP1 is a well-established biomarker that captures aspects of the inflammatory response. Similarly, IL18, which is a pro-inflammatory cytokine involved in a range of inflammatory responses including pyroptosis^60^, has been found to be increased in both blood serum and the superior frontal cortex at postmortem in people with FTD^30^. Higher CSF levels of IL18 were also detected in fast-progressing ALS with a C9orf72 mutation^61^. Several proteins with high loadings onto the protein canonical variate, including MMP10, CCL25, LAP-TGF-β1, OPG, and SIRT2, have received little to no attention in FTD fluid biomarkers research. The convergence of these novel biomarkers onto the canonical variate implicates extracellular matrix remodelling, immune cell trafficking, and osteoimmune signalling as underexplored pathological processes in FTD pathogenesis, warranting further investigation.

Although TNF-*α* and IL-6 were not among the top-loading CSF proteins in our rCCA, this may reflect cohort, tissue or platform differences rather than a lack of biological relevance. Previous research has linked increased plasma levels of TNF-*α* and IL-6 to accelerated longitudinal atrophy in the functional and temporal lobes in MAPT mutation carriers, while in C9orf72 mutation carriers higher TNF-*α* abundance was as-sociated with faster functional and cognitive deterioration^31^. Given that survival pro-vides a more objective quantification of disease progression than certain clinical scales, Malpetti et al (2024)^23^ implemented a data-driven approach to capture a pro-inflammatory signature driven primarily by TNF-*α*, TNF-R1, M-CSF, IL-17A, IL-12, IP-10 and IL-6. This pro-inflammatory pattern was significantly associated with a higher probability of shorter survival in sporadic FTD, as well as higher levels of brain inflammation^23^. Linking peripheral immune signalling to in-vivo molecular brain imaging, utilising the [^11^C]-PK11195 radiotracer (a proxy measure for neuroinflammation, binding TSPO^62^ ^63^), previous PET studied showed elevated neuroinflammation in frontotemporal regions of symptomatic carriers of FTD-related gene mutations as compared to controls^64^. However, the pattern of increased [^11^C]-PK11195 signal was heterogeneous within the symptomatic FTD mutation-carrier group, closely matching brain regions that relate to their clinical syndrome; for example, participants with a MAPT mutation with a clinical diagnosis of bvFTD had elevated binding in fronto-striatal regions, whereas a participant with PPA exhibited elevated signal in temporal regions. A single case study has also described elevated [^11^C]-PK11195 signal in a presymptomatic MAPT carriers, in absence of detectable regional atrophy^65^. However, capturing the complex and intertwined links between inflammatory signalling and localised atrophy patterns requires robust multimodal statistical integration. Our implemented rCCA model addresses this complexity.

We deployed an rCCA, which is a model that captures shared variances between our two data sets (protein and MRI). To examine whether a non-brain-related inflammatory pattern is also clinically relevant we residualised our proteins against protein-CV and extracted the residuals. This effectively removed any variance that is captured within the inflammation-atrophy axis. Applying a PCA to the extracted residuals we derived an inflammation pattern that was completely decoupled from the atrophy pattern. Interestingly, this inflammation signature was not linked to any of the clinical outcomes within the symptomatic group. This suggests that a generalised immune pattern was not sufficient to explain clinical decline in our cohort, whereas the inflammatory protein-brain axis was able to predict clinical decline. Hence, the shared variance between the protein and MRI data is clinically relevant and inter-twined. However, CXCL9 levels (the most highly loaded protein derived from the rCCA) alone were associated with clinical progression in participants who phenoconverted. This supplements the findings from Asken et al. (2023)^31^, which identified TNF-*α* and IL6 as clinically relevant inflammatory markers in genetic FTD. Further research is needed to investigate whether stepwise therapeutic strategies utilising compounds targeting different parts of the immune system could be efficient disease-modifying interventions in people with FTD.

The inflammation-atrophy axis identified in symptomatic participants has also translational relevance both as a therapeutic target and as a multivariate biomarker component for trial design. Three top-loading proteins map onto pathways with existing pharmacological tool compounds: MCP1/CCL2, IL-18 and CCL3. These can be considered in terms of translational and repurposing readiness from other fields into FTD trials. MCP1 has been implicated in HIV-associated neurocognitive impairment, which affects around 30-50% of people living with HIV. ^88^ ^89^ In a small single-arm study, the dual CCR2/CCR5 antagonist cenicriviroc was associated with improved cognitive performance and reduced inflammation over 24 weeks, although it has not yet been tested in FTD. ^90^ The NLRP3 inflammasome-caspase-1 IL18 axis is a key mediator of innate immune activation across multiple diseases and can be targeted via IL-18 neutralisation or NLRP3/caspase-1 inhibition; notably, the oral NLRP3 in-hibitor dapansutrile is in Phase 2 testing in early Parkinson’s disease (DAPA-PD; NCT07157735)^91^ ^92^ ^93^ ^94^ ^95^. CCL3 is a chemotactic cytokine signalling through CCR1/CCR5. ^69^ Maraviroc, a CCR5 antagonist approved for HIV, has been shown by Festa et al (2023) to rescue autophagic and neurodegenerative phenotypes in mouse models^96^ ^97^ and can cross the blood-brain barrier in people with HIV^66^. Collectively, these observations suggest that targeting MCP1/CCL2, IL-18 and CCL3 biology may hold therapeutic promise in FTD and related dementias. Because the atrophy-related inflammatory pattern we identified was transdiagnostic, CSF inflammatory panels are unlikely to offer strong differential-diagnostic value across the FTD continuum. However, this same transdiagnostic quality suggests an integrated immunotherapeutic strategy may prove beneficial across individual FTD-spectrum diseases, rather than requiring disease-specific approaches. This is further supported by a growing body of clinical evidence linking inflammatory profiles to structural brain changes and shorter survival.

Several studies have demonstrated early brain changes in genetic FTD occurring within the presymptomatic window^67^ ^14^. However, in the present study, the CCA-defined inflammatory protein component and the inflammation-related atrophy profiles revealed no statistically significant differences between healthy controls and presymptomatic mutation carriers. This is consistent with De Houwer et al. (2025)^28^, who employed proximity extension assay Olink technology to quantify over 2,900 proteins in CSF samples of people with genetic FTD and similarly identified no differentially abundant proteins between presymptomatic participants and controls. One explanation is that CSF sampling in our cohort occurred early in the disease trajectory, at a point when pathological processes remain below the detection threshold of current proteomic tools. Had samples been collected closer to phenoconversion, differences in both inflammatory proteins and inflammation-related MRI patterns relative to controls would more likely be apparent. To address this, we ran sensitivity analyses re-stratifying our participants by CDR-SB scores, which more sensitively capture granular clinical changes than the simple absence of a formal clinical diagnosis. We identified stepwise (although not statistically significant) divergence between the prodromal and asymptomatic stages in the MRI-CV and protein-CV scores, as well as at the individual protein levels of CXCL9 and NfL. Future studies should also explore whether composite scores derived from integrating CSF proteomic and MRI data could enhance clinical trials as stratification or endpoint tools in early phases of FTD.

Another aspect to consider is the limitations of the proteomic approach. Although proximity extension assay technology robustly quantifies a pre-specified set of proteins, the human proteome encompasses tens of thousands of distinct proteins, the majority of which remain unmeasured^68^ ^69^ ^70^. Crucially, this platform requires intact proteins with accessible epitopes for dual-antibody binding, and therefore cannot reliably detect short peptides, protein fragments, or heavily modified proteoforms - species that may undergo early changes in FTD and carry prognostic or mechanistic significance. Unbiased, mass spectrometry-based proteomics approaches therefore offer a complementary and potentially more sensitive strategy for biomarker discovery in this context. In line with this, a recent GENFI study^16^, employing mass spectrometry in CSF of participants with genetic FTD, found that the immune response co-expression module did not significantly differentiate presymptomatic carriers from controls, though other proteomic clusters showed presymptomatic changes, providing further evidence that immune dysregulation may not be detectable in CSF during the presymptomatic phase. Given that neither CSF inflammation proteomics nor the CCA-derived profiles detected significant immune dysregulation in presymptomatic carriers, it is possible that neuroinflammatory activation and immunological changes may be spatially restricted to the brain parenchyma at this disease stage and not yet reflected in the CSF compartment. Future studies incorporating positron emission tomography with translocator protein (TSPO) radioligands could directly visualise microglial-driven neuroinflammation in presymptomatic mutation carriers^71^, offering a more sensitive and spatially resolved window into the temporal dynamics of neuroin-flammation in presymptomatic FTD.

Several limitations of this study should be considered when interpreting our findings. First, whilst the Olink Target 96 Inflammation platform is scalable and widely accessible, it quantifies 92 pre-selected inflammatory proteins, but does not capture the whole complexity of inflammatory cascades and other disease-relevant pathways, including synaptic and metabolic changes. Second, medical history data on past or current infections and inflammatory conditions were not available. However, our study focussed on a atrophy-related inflammatory protein pattern in CSF, which may reduce capturing signals related to peripheral conditions. Third, our study was insufficiently powered to investigate group differences in the inflammation-atrophy axis between gene mutation cohorts (C9orf72, GRN, or MAPT). Pooling the genetic sub-groups in the rCCA may mask mutation-specific inflammation-atrophy associations, as previous studies identified mutation-specific differences in protein abundance^28^ ^27^. Finally, CSF proteomic data were available cross-sectionally only and the rCCA deployed is a linear statistical model that assumes linear patterns in the protein-brain covariance matrix; hence, non-linear associations are not captured. This is particularly relevant for inflammation, which is not a static process but rather a highly complex, dynamic mechanism that may change over time. Future research incorporating longitudinal data is needed to capture non-linear trajectories of inflammatory processes across the pre-symptomatic and symptomatic windows of FTD, as well as multimodal and sensitive approaches that can capture changes in brain *versus* peripheral inflammation early in FTD, even before symptoms.

### Conclusion

In conclusion, our study identified a robust data-driven association between CSF inflammatory proteins and brain atrophy in genetic FTD, that is driven predominantly by older symptomatic participants. The atrophy-related inflammatory protein pattern identified in this study is clinically relevant, predicting both disease progression and cross-sectional executive dysfunction in symptomatic individuals. These findings support the use of multivariate approaches to examine complex relationships and dynamics between brain atrophy, inflammation and clinical outcomes in FTD. Our study provides further evidence that inflammatory pathways are implicated in genetic FTD and related to worse clinical outcomes, representing potential therapeutic targets in this population.

## Data Availability

All data produced in the present study are available upon reasonable request to the authors

## Acknowledgements

We thank participant volunteers and their families for their participation in this study. For the purpose of open access, the authors have applied a Creative Commons At-tribution (CC BY) license to any Author Accepted Manuscript version arising from this submission. This work is licensed under a Creative Commons Attribution 4.0 International License.

---

| Author | Affiliation |
|---|---|
| Amelia Ble-sius | Department of Neurodegenerative Disease, Dementia Research Centre, UCL Queen Square Institute of Neurology, London, UK |
| David Cash | Department of Neurodegenerative Disease, Dementia Research Centre, UCL Queen Square Institute of Neurology, London, UK |
| Clementine Cheyne | Department of Neurodegenerative Disease, Dementia Research Centre, UCL Queen Square Institute of Neurology, London, UK |
| Sophie Gold-smith | Department of Neurodegenerative Disease, Dementia Research Centre, UCL Queen Square Institute of Neurology, London, UK |
| Luna Norden-strom | Department of Neurodegenerative Disease, Dementia Research Centre, UCL Queen Square Institute of Neurology, London, UK |
| Victoria Pau-wels Romero | Department of Neurodegenerative Disease, Dementia Research Centre, UCL Queen Square Institute of Neurology, London, UK |
| Hannah Rosenheim | Department of Neurodegenerative Disease, Dementia Research Centre, UCL Queen Square Institute of Neurology, London, UK |
| David L. Tho-mas | Neuroimaging Analysis Centre, Department of Brain Repair and Rehabilitation, UCL Institute of Neurology, Queen Square, London, UK |
| Maura Malpetti | Department of Clinical Neurosciences, University of Cambridge, Cambridge, UK |
| Alexander Murley | Department of Clinical Neurosciences, University of Cambridge, Cambridge, UK |
| Sean Tan | Department of Clinical Neurosciences, University of Cambridge, Cambridge, UK |
| Sonia Bellini | Molecular Markers Laboratory, IRCCS Istituto Centro San Giovanni di Dio Fatebenefratelli, Brescia, Italy |
| Valentina Cantoni | Department of Clinical and Experimental Sciences, University of Brescia, Brescia, Italy |
| Roberto Gas-parotti | Neuroradiology Unit, University of Brescia, Brescia, Italy |
| Roberta Ghi-doni | Molecular Markers Laboratory, IRCCS Istituto Centro San Giovanni di Dio Fatebenefratelli, Brescia, Italy |
| Enrico Premi | Stroke Unit, ASST Brescia Hospital, Brescia, Italy |
| Andrea Arighi | Neurodegenerative Diseases Unit, Fondazione IRCCS Ca' Granda Ospedale Maggiore Policlinico, Milan, Italy |
| Vittoria Bor-racci | Neurodegenerative Diseases Unit, Fondazione IRCCS Ca' Granda, Ospedale Maggiore Policlinico, Milan, Italy |
| Chiara Fenoglio | Department of Biomedical, Surgical and Dental Sciences, University of Milan, and Fondazione IRCCS Ca' Granda, Ospedale Maggiore Policlinico, Milan, Italy |
| Manuela Pin-tus | Neurodegenerative Diseases Unit, Fondazione IRCCS Ca' Granda, Ospedale Maggiore Policlinico, Milan, Italy |
| Emanuela Rotondo | Neurodegenerative Diseases Unit, Fondazione IRCCS Ca' Granda, Ospedale Maggiore Policlinico, Milan, Italy |
| Luca Sacchi | Neurodegenerative Diseases Unit, Fondazione IRCCS Ca' Granda, Ospedale Maggiore Policlinico, Milan, Italy |
| Maria Ser-pente | Neurodegenerative Diseases Unit, Fondazione IRCCS Ca' Granda Ospedale Maggiore Policlinico, Milan, Italy |
| Vittoria Aprea | Fondazione IRCCS Istituto Neurologico Carlo Besta, Milan, Italy |
| Paola Caroppo | Fondazione IRCCS Istituto Neurologico Carlo Besta, Milan, Italy |
| Marcella Ca-tania | Fondazione IRCCS Istituto Neurologico Carlo Besta, Milan, Italy |
| Giorgio Ge-losa | Fondazione IRCCS Istituto Neurologico Carlo Besta, Milan, Italy |
| Gemma Lombardi | Fondazione IRCCS Istituto Neurologico Carlo Besta, Milan, Italy |
| Pietro TIraboschi | Fondazione IRCCS Istituto Neurologico Carlo Besta, Milan, Italy |
| Sandra Black | Sunnybrook Health Sciences Centre, Sunnybrook Research Institute, University of Toronto, Toronto, Canada |
| Morris Freed-man | Baycrest Health Sciences, Rotman Research Institute, University of Toronto, Toronto, Canada |
| Sara Mitchell | Sunnybrook Health Sciences Centre, Sunnybrook Research Institute, University of Toronto, Toronto, Canada |
| Ekaterina Ro-gaeva | Tanz Centre for Research in Neurodegenerative Diseases, University of Toronto, Toronto, Can-ada |
| David Tang-Wai | The University Health Network, Krembil Research Institute, Toronto, Canada |
| Olaiya Aro | Department of Neurology, Erasmus Medical Centre, Rotterdam, Netherlands |
| Liset de Boer | Department of Neurology, Erasmus Medical Centre, Rotterdam, Netherlands |
| Julie De Hou-wer | Department of Neurology, Erasmus Medical Centre, Rotterdam, Netherlands |
| Laura Donker Kaat | Department of Neurology, Erasmus Medical Centre, Rotterdam, Netherlands |
| Elise Dopper | Department of Neurology, Erasmus Medical Centre, Rotterdam, Netherlands |
| Jackie Poos | Department of Neurology, Erasmus Medical Centre, Rotterdam, Netherlands |
| Babette Reichard | Department of Neurology, Erasmus Medical Centre, Rotterdam, Netherlands |
| Tine Swarten-broekx | Department of Neurology, Erasmus Medical Centre, Rotterdam, Netherlands |
| Valentina Bessi | Department of Neuroscience, Psychology, Drug Research and Child Health, Florence, Italy |
| Stefano Chiti | Neuroradiology Unit, Department of Experimental and Clinical Biomedical Sciences, University of Florence, Florence, Italy |
| Giulia Gia-comucci | Department of Neuroscience, Psychology, Drug Research and Child Health, University of Flor-ence, Florence, Italy |
| Andrea Gine-stroni | Neuroradiology Unit, Department of Radiology, Careggi University Hospital, Florence, Italy |
| Valentina Mo-schini | SOD Neurologia Dipartimento di Neuroscienze degli Organi di Senso, AOU Careggi, Florence, Italy |
| Sonia Padigli-oni | SOD Neurologia Dipartimento di Neuroscienze degli Organi di Senso, AOU Careggi, Florence, Italy |
| Andrew Doug-las | Nuffield Department of Clinical Neurosciences, University of Oxford |
| Kevin Talbot | Nuffield Department of Clinical Neurosciences, University of Oxford |
| Martin Turner | Nuffield Department of Clinical Neurosciences, University of Oxford |
| Vesna Jelic | Department of Neurobiology, Care Sciences and Society; Division of Clinical Geriatrics, Karolin-ska Institutet, Stockholm, Sweden |
| Elena Rodri-guez-Vieitez | Department of Neurobiology, Care Sciences and Society, Center for Alzheimer Research, Divi-sion of Neurogeriatrics, Bioclinicum, Karolinska Institutet, Solna, Sweden |
| Melissa Taheri Rydell | Department of Neurobiology, Care Sciences and Society, Center for Alzheimer Research, Divi-sion of Neurogeriatrics, , Bioclinicum, Karolinska Institutet, Solna, Sweden |
| Henrik Viklund | Theme Inflammation and aging, Karolinska University Hospital, Sweden |
| Caroline Dal-laire-Théroux | Clinique Interdisciplinaire de Mémoire, CHU de Québec, and CRCHU de Québec, Université Laval, Québec, Canada |
| Anna Antonell | Alzheimer's and other cognitive disorders unit. Hospital Clínic de Barcelona. FRCB-IDIBAPS. University of Barcelona. CIBERNED, Barcelona, Spain |
| Mircea Balasa | Alzheimer's and other cognitive disorders unit. Hospital Clínic de Barcelona. FRCB-IDIBAPS. University of Barcelona. CIBERNED, Barcelona, Spain |
| Sergi Borrego-Ecija | Alzheimer's and other cognitive disorders unit. Hospital Clínic de Barcelona. FRCB-IDIBAPS. University of Barcelona. CIBERNED, Barcelona, Spain |
| Paula Ca-nasto | Alzheimer's and other cognitive disorders unit. Hospital Clínic de Barcelona. FRCB-IDIBAPS. University of Barcelona. CIBERNED, Barcelona, Spain |
| Jordi Junca | Alzheimer's and other cognitive disorders unit. Hospital Clínic de Barcelona. FRCB-IDIBAPS. University of Barcelona. CIBERNED, Barcelona, Spain |
| Albert Lladó | Alzheimer's and other cognitive disorders unit. Hospital Clínic de Barcelona. FRCB-IDIBAPS. University of Barcelona. CIBERNED, Barcelona, Spain |
| Tiago Costa-Coelho | Faculty of Medicine, University of Lisbon, Lisbon, Portugal |
| Carolina Ma-ruta | Laboratory of Language Research, Centro de Estudos Egas Moniz, Faculty of Medicine, Uni-versity of Lisbon, Lisbon, Portugal |
| Gabriel Miltenberger | Faculty of Medicine, University of Lisbon, Lisbon, Portugal |
| Ana Verdelho | Faculty of Medicine, University of Lisbon, Lisbon, Portugal |
| Patricia Alves | Biogipuzkoa Health Research Institute, Neurosciences Area, Group of Neurodegenerative Dis-eases, 20014, San Sebastian, Spain |
| Myriam Barandiaran | Cognitive Disorders Unit, Department of Neurology, Donostia University Hospital, San Sebas-tian, Gipuzkoa, Spain |
| Ioana Croitoru | Biogipuzkoa Health Research Institute, Neurosciences Area, Group of Neurodegenerative Dis-eases, 20014, San Sebastian, Spain |
| Alazne Ga-bilondo | Cognitive Disorders Unit, Department of Neurology, Donostia University Hospital, San Sebas-tian, Gipuzkoa, Spain |
| Iliana Qui-ñones | Biogipuzkoa Health Research Institute, Neurosciences Area, Group of Neurodegenerative Dis-eases, 20014, San Sebastian, Spain |
| Mikel Tainta | Cognitive Disorders Unit, Department of Neurology, Donostia University Hospital, San Sebas-tian, Gipuzkoa, Spain |
| Lukas Bei-chert | Division of Translational Genomics of Neurodegenerative Diseases, Tübingen, Germany |
| Benjamin Bender | Department of Diagnostic and Interventional Neuroradiology, Tübingen, Germany |
| Lisa Graf | Division of Translational Genomics of Neurodegenerative Diseases, Tübingen, Germany |
| *Mathieu Van-denbulcke* | *Geriatric Psychiatry Service, University Hospitals Leuven, Leuven, Belgium* |
| *Koen Poesen* | *Laboratory for Molecular Neurobiomarker Research, KU Leuven, Leuven, Belgium* |
| *Sien Van Daele* | *Department of Human Genetics, Leuven, Belgium* |
| *Aline Delva* | *Neurology service, Leuven, Belgium* |
| Maxime Mon-tembeault | Douglas Research Center, Department of Psychiatry, McGill University, Montreal, Canada |
| Guy Rouleau | Montreal Neurological Institute, Department of Neurology & Neurosurgery, McGill University, Montreal, Canada |
| Pierre-François Pra-dat | Reference Center for ALS, Département de Neurologie, AP-HP - Hôpital Pitié-Salpêtrière, Paris, France |
| Giorgia Querin | Paris Brain Institute - Institut du Cerveau - ICM, Sorbonne Université, Inserm U1127, CNRS UMR 7225, AP-HP - Hôpital Pitié-Salpêtrière, Paris, France |
| Daisy Rinaldi | Paris Brain Institute - Institut du Cerveau - ICM, Sorbonne Université, Inserm U1127, CNRS UMR 7225, AP-HP - Hôpital Pitié-Salpêtrière, Paris, France |
| Anna Stock-bauer | Neurologische Klinik, Ludwig-Maximilians-Universität München, Munich, Germany |
| Alexander Bernhardt | Neurologische Klinik, Ludwig-Maximilians-Universität München, Munich, Germany |
| Ellen Merli | Neurologische Klinik, Ludwig-Maximilians-Universität München, Munich, Germany |
| Elisabeth Wlasich | Neurologische Klinik, Ludwig-Maximilians-Universität München, Munich, Germany |
| Georg Nübling | Neurologische Klinik, Ludwig-Maximilians-Universität München, Munich, Germany |
| Julia Kuster-mann | Neurologische Klinik, Ludwig-Maximilians-Universität München, Munich, Germany |
| Olivia Wage-mann | Neurologische Klinik, Ludwig-Maximilians-Universität München, Munich, Germany |
| Sonja Schönecker | Neurologische Klinik, Ludwig-Maximilians-Universität München, Munich, Germany |
| Lena Marth | Neurologische Klinik, Ludwig-Maximilians-Universität München, Munich, Germany |
| *Sarah Anderl-Straub* | *Department of Neurology, University of Ulm, Ulm, Germany* |
| Ilse Dewa-chter | Department of Neurosciences, Biomedical Research Institute BIOMED, Hasselt University, Hasselt, Belgium |
| Stéphanie De Keulenaer | Department of Biomedical Sciences, University of Antwerp, Antwerp, Belgium |
| Rosa Rade-makers | VIB Center for Molecular Neurology, VIB, Antwerp, Belgium |
| Marijne Vandebergh | VIB Center for Molecular Neurology, VIB, Antwerp, Belgium |
| Thijs Vande Vyvere | Department of Radiology, University Hospital of Antwerp (UZA), Antwerp, Belgium |
| Sara Van Mossevelde | Department of Neurology, University Hospital of Antwerp (UZA), Antwerp, Belgium |
| Xavier Del-beuck | Univ. Lille, Inserm, CHU Lille, Lille Neuroscience & Cognition, LabEx DISTALZ, F-59000, Lille, France |
| Agnès Denève | Univ. Lille, Inserm, CHU Lille, Lille Neuroscience & Cognition, LabEx DISTALZ, F-59000, Lille, France |
| Simon Lecerf | Univ. Lille, Inserm, CHU Lille, Lille Neuroscience & Cognition, LabEx DISTALZ, F-59000, Lille, France |
| Juhana Ha-kumäki | University of Eastern Finland, Kuopio, Finland and Kuopio University Hospital, Kuopio, Finland |
| Sanna Han-nonen | University of Eastern Finland, Kuopio, Finland and Kuopio University Hospital, Kuopio, Finland |
| Juho Kalapu-das | University of Eastern Finland, Kuopio, Finland and Kuopio University Hospital, Kuopio, Finland |
| Annemari Kilpeläinen | University of Eastern Finland, Kuopio, Finland and Kuopio University Hospital, Kuopio, Finland |
| Ave Kivisild | Kuopio University Hospital, Kuopio, Finland |
| Mervi Könönen | University of Eastern Finland, Kuopio, Finland and Kuopio University Hospital, Kuopio, Finland |
| Inês Baldeiras | Neurochemistry Laboratory, Coimbra University Hospital, University of Coimbra, Coimbra, Por-tugal |
| Miguel Cas-telo-Branco | Institute of Nuclear Sciences Applied to Health (ICNAS), University of Coimbra, Coimbra, Por-tugal |
| João Durães | Neurology Department, Centro Hospitalar e Universitario de Coimbra, Coimbra, Portugal |
| Marisa Lima | Neurology Department, Centro Hospitalar e Universitario de Coimbra, Coimbra, Portugal |
| Maria Rosario Almeida | Centre of Neurosciences and Cell Biology, University of Coimbra, Coimbra, Portugal |
| Miguel Tábuas-Pereira | Neurology Department, Centro Hospitalar e Universitario de Coimbra, Coimbra, Portugal |
| Annerose Engel | Cognitive Neurology Clinic, University Hospital Leipzig, Germany |
| Maryna Polyakova | Department Neurology, Max Planck Institute for Human Cognitive and Brain Sciences, Leipzig, Germany |
| Frank Regen-brecht | Cognitive Neurology Clinic, University Hospital Leipzig, Germany |
| Angelika Thöne-Otto | Cognitive Neurology Clinic, University Hospital Leipzig, Germany |
| Anna-Leena Heikkinen | Neurocenter, Neurology, Oulu University Hospital, Oulu, Finland and Research Unit of Clinical Medicine, Neurology, University of Oulu, Finland |
| Vesa Kiviniemi | Oulu Functional Neuroimaging, Research Unit of Health Sciences and Technology, University of Oulu, Oulu, Finland |
| Vesa Korho-nen | Oulu Functional Neuroimaging, Research Unit of Health Sciences and Technology, University of Oulu, Oulu, Finland |
| Noora-Maria Suhonen | Neurocenter, Neurology, Oulu University Hospital, Oulu, Finland and Research Unit of Clinical Medicine, Neurology, University of Oulu, Finland |
| Laura Göschel | Department of Neurology; Universitätsmedizin Berlin, Berlin, Germany |
| Jonah Nietiet | Department of Neurology; Universitätsmedizin Berlin, Berlin, Germany |
| Anna Söllner | Department of Neurology; Universitätsmedizin Berlin, Berlin, Germany |
| Teresa Dell'Abbate | Center for Neurodegenerative Diseases and the Aging Brain. University of Bari "Aldo Moro" at Pia Fondazione, Tricase, Italy |
| Daniele Urso | Department of Clinical Research in Neurology, University of Bari, Italy |
| Daniel Alcolea | Department of Neurology, Hospital de la Santa Creu i Sant Pau, Barcelona, Spain |
| Sara Bernal | Department of Genetics, Hospital de la Santa Creu i Sant Pau, Barcelona, Spain |
| Oriol Dols | Department of Neurology, Hospital de la Santa Creu i Sant Pau, Barcelona, Spain |
| Juan Fortea | Department of Neurology, Hospital de la Santa Creu i Sant Pau, Barcelona, Spain |
| Jesus García-Castro | Department of Neurology, Hospital de la Santa Creu i Sant Pau, Barcelona, Spain |
| Alberto Lleó | Department of Neurology, Hospital de la Santa Creu i Sant Pau, Barcelona, Spain |
| Sara Rubio-Guerra | Department of Neurology, Hospital de la Santa Creu i Sant Pau, Barcelona, Spain |
| Judit Selma-González | Department of Neurology, Hospital de la Santa Creu i Sant Pau, Barcelona, Spain |

## References

1. Gorno-Tempini ML, Hillis AE, Weintraub S, et al. Classification of primary progressive aphasia and its variants. Neurology. 2011;76(11):1006–1014. doi:10.1212/WNL.0b013e31821103e6

2. Rascovsky K, Hodges JR, Knopman D, et al. Sensitivity of revised diagnostic criteria for the behavioural variant of frontotemporal dementia. Brain. 2011;134(9):2456–2477. doi:10.1093/brain/awr179

3. Grossman M, Seeley WW, Boxer AL, et al. Frontotemporal lobar degeneration. Nat Rev Dis Primers. 2023;9(1):40. doi:10.1038/s41572-023-00447-0

4. DeJesus-Hernandez M, Mackenzie IR, Boeve BF, et al. Expanded GGGGCC hexanucleotide repeat in noncoding region of C9ORF72 causes chromosome 9p-linked FTD and ALS. Neuron. 2011;72(2):245–256. doi:10.1016/j.neuron.2011.09.011

5. Renton AE, Majounie E, Waite A, et al. A hexanucleotide repeat expansion in C9ORF72 is the cause of chromosome 9p21-linked ALS-FTD. Neuron. 2011;72(2):257–268. doi:10.1016/j.neuron.2011.09.010

6. Baker M, Mackenzie IR, Pickering-Brown SM, et al. Mutations in progranulin cause tau-negative frontotemporal dementia linked to chromosome 17. Nature. 2006;442(7105):916–919. doi:10.1038/nature05016

7. Cruts M, Gijselinck I, van der Zee J, et al. Null mutations in progranulin cause ubiquitin-positive frontotemporal dementia linked to chromosome 17q21. Nature. 2006;442(7105):920–924. doi:10.1038/nature05017

8. Hutton M, Lendon CL, Rizzu P, et al. Association of missense and 5′-splice-site mutations in tau with the inherited dementia FTDP-17. Nature. 1998;393(6686):702–705. doi:10.1038/31508

9. Lashley T, Rohrer JD, Mead S, Revesz T. Review: an update on clinical, genetic and pathological aspects of frontotemporal lobar degenerations. Neuropathol Appl Neurobiol. 2015;41(7):858–881. doi:10.1111/nan.12250

10. Rohrer JD, Nicholas JM, Cash DM, et al. Presymptomatic cognitive and neuroanatomical changes in genetic frontotemporal dementia in the Genetic Fronto-temporal dementia Initiative (GENFI) study: a cross-sectional analysis. The Lancet Neurology. 2015;14(3):253–262. doi:10.1016/S1474-4422(14)70324-2

11. Malpetti M, Holland N, Jones PS, et al. Synaptic density in carriers of C9orf72 mutations: a [11C]UCB-J PET study. Ann Clin Transl Neurol. 2021;8(7):1515–1523. doi:10.1002/acn3.51407

12. Tsvetanov KA, Malpetti M, Jones PS, et al. Cellular signatures underlying functional resilience in presymptomatic frontotemporal dementia. Brain. Published online November 24, 2025:awaf443. doi:10.1093/brain/awaf443

13. Staffaroni AM, Bajorek L, Casaletto KB, et al. Assessment of executive function declines in presymptomatic and mildly symptomatic familial frontotemporal dementia: NIH-EXAMINER as a potential clinical trial endpoint. Alzheimer’s & Dementia. 2020;16(1):11–21. doi:10.1016/j.jalz.2019.01.012

14. Malpetti M, Jones PS, Tsvetanov KA, et al. Apathy in presymptomatic genetic frontotemporal dementia predicts cognitive decline and is driven by structural brain changes. Alzheimer’s & Dementia. 2021;17(6):969–983. doi:10.1002/alz.12252

15. Bergström S, Öijerstedt L, Remnestål J, et al. A panel of CSF proteins separates genetic frontotemporal dementia from presymptomatic mutation carriers: a GENFI study. Mol Neurodegeneration. 2021;16(1):79. doi:10.1186/s13024-021-00499-4

16. Sogorb-Esteve A, Weiner S, Simrén J, et al. Proteomic analysis reveals distinct cerebrospinal fluid signatures across genetic frontotemporal dementia sub-types. Sci Transl Med. 2025;17(784):eadm9654. doi:10.1126/scitranslmed.adm9654

17. Bright F, Werry EL, Dobson-Stone C, et al. Neuroinflammation in frontotempo-ral dementia. Nat Rev Neurol. 2019;15(9):540–555. doi:10.1038/s41582-019-0231-z

18. Malpetti M, Cope TE, Street D, et al. Microglial activation in the frontal cortex predicts cognitive decline in frontotemporal dementia. Brain. 2023;146(8):3221–3231. doi:10.1093/brain/awad078

19. Katisko K, Solje E, Korhonen P, et al. Peripheral inflammatory markers and clinical correlations in patients with frontotemporal lobar degeneration with and without the C9orf72 repeat expansion. J Neurol. 2020;267(1):76–86. doi:10.1007/s00415-019-09552-1

20. McCauley ME, Baloh RH. Inflammation in ALS/FTD pathogenesis. Acta Neuropathol. 2019;137(5):715–730. doi:10.1007/s00401-018-1933-9

21. Erichsen PA, Henriksen EE, Nielsen JE, Ejlerskov P, Simonsen AH, Toft A. Immunological Fluid Biomarkers in Frontotemporal Dementia: A Systematic Re-view. Biomolecules. 2025;15(4):473. doi:10.3390/biom15040473

22. Strauss A, Swann P, Kigar SL, et al. Peripheral innate immunophenotype in neurodegenerative disease: blood-based profiles and links to survival. Mol Psychiatry. Published online October 29, 2024. doi:10.1038/s41380-024-02809-w

23. Malpetti M, Swann P, Tsvetanov KA, et al. Blood inflammation relates to neuroinflammation and survival in frontotemporal lobar degeneration. Brain. 2025;148(2):493–505. doi:10.1093/brain/awae269

24. Bossù P, Salani F, Alberici A, et al. Loss of function mutations in the progranulin gene are related to pro-inflammatory cytokine dysregulation in frontotemporal lobar degeneration patients. J Neuroinflammation. 2011;8(1):65. doi:10.1186/1742-2094-8-65

25. Sjogren M. Increased intrathecal inflammatory activity in frontotemporal dementia: pathophysiological implications. Journal of Neurology, Neurosurgery & Psychiatry. 2004;75(8):1107–1111. doi:10.1136/jnnp.2003.019422

26. Miller ZA, Rankin KP, Graff-Radford NR, et al. TDP-43 frontotemporal lobar degeneration and autoimmune disease. Journal of Neurology, Neurosurgery & Psychiatry. 2013;84(9):956–962. doi:10.1136/jnnp-2012-304644

27. Fenoglio C, Serpente M, Arcaro M, et al. Inflammatory plasma profile in genetic symptomatic and presymptomatic Frontotemporal Dementia − A GENFI study. Brain, Behavior, and Immunity. 2024;122:231–240. doi:10.1016/j.bbi.2024.08.030

28. De Houwer JFH, Dopper EG, Van Buuren R, et al. Distinct proteomic CSF profiles in genetic frontotemporal lobar degeneration. Brain. Published online De-cember 4, 2025:awaf457. doi:10.1093/brain/awaf457

29. Gamez N, Eid AM, Pascual B, et al. Peripheral immune profiling in frontotem-poral dementia. Brain Communications. Published online March 13, 2026:fcag089. doi:10.1093/braincomms/fcag089

30. Lok HC, Katzeff JS, Hodges JR, et al. Elevated GRO-α and IL-18 in serum and brain implicate the NLRP3 inflammasome in frontotemporal dementia. Sci Rep. 2023;13(1):8942. doi:10.1038/s41598-023-35945-4

31. Asken BM, Ljubenkov PA, Staffaroni AM, et al. Plasma inflammation for predicting phenotypic conversion and clinical progression of autosomal dominant frontotemporal lobar degeneration. J Neurol Neurosurg Psychiatry. 2023;94(7):541–549. doi:10.1136/jnnp-2022-330866

32. Ullgren A, Rydell MT, Bergström S, et al. CSF protein biomarkers are associated with atrophy and symptom severity in genetic FTD: a GENFI study. J Neural Transm. Published online February 19, 2026. doi:10.1007/s00702-026-03118-y

33. Assogna M, Di Lorenzo F, Bonnì S, et al. Phase 2 study of palmitoylethanolamide combined with luteoline in frontotemporal dementia patients. Brain Communications. 2025;7(2):fcaf080. doi:10.1093/braincomms/fcaf080

34. Koch G, Assogna M, Gadola Y, et al. Safety and efficacy of rotigotine in patients with frontotemporal dementia: a phase 2, double-blind, randomized, placebo-controlled, multicenter trial. The Lancet Regional Health - Europe. 2025;57:101409. doi:10.1016/j.lanepe.2025.101409

35. Morris JC. The Clinical Dementia Rating (CDR): Current version and scoring rules. Neurology. 1993;43(11):2412. doi:10.1212/WNL.43.11.2412-a

36. Miyagawa T, Brushaber D, Syrjanen J, et al. Use of the CDR® plus NACC FTLD in mild FTLD: Data from the ARTFL/LEFFTDS consortium. Alzheimer’s Dement. 2020;16(1):79–90. doi:10.1016/j.jalz.2019.05.013

37. Knopman DS, Weintraub S, Pankratz VS. Language and behavior domains enhance the value of the clinical dementia rating scale. Alzheimer’s & Dementia. 2011;7(3):293–299. doi:10.1016/j.jalz.2010.12.006

38. Peakman G, Russell LL, Convery RS, et al. Comparison of clinical rating scales in genetic frontotemporal dementia within the GENFI cohort. J Neurol Neurosurg Psychiatry. 2022;93(2):158–168. doi:10.1136/jnnp-2021-326868

39. Wechsler D. WAIS-R: Manual: Wechsler adult intelligence scale-revised. Published online 1981.

40. Bocchetta M, Todd EG, Peakman G, et al. Differential early subcortical involvement in genetic FTD within the GENFI cohort. NeuroImage: Clinical. 2021;30:102646. doi:10.1016/j.nicl.2021.102646

41. Passamonti L, Tsvetanov KA, Jones PS, et al. Neuroinflammation and Functional Connectivity in Alzheimer’s Disease: Interactive Influences on Cognitive Performance. J Neurosci. 2019;39(36):7218–7226. doi:10.1523/JNEUROSCI.2574-18.2019

42. Tsvetanov KA, Henson RNA, Tyler LK, et al. Extrinsic and Intrinsic Brain Network Connectivity Maintains Cognition across the Lifespan Despite Accelerated Decay of Regional Brain Activation. J Neurosci. 2016;36(11):3115–3126. doi:10.1523/JNEUROSCI.2733-15.2016

43. Tsvetanov KA, Spindler LRB, Stamatakis EA, et al. Hospitalisation for COVID-19 predicts long lasting cerebrovascular impairment: A prospective observational cohort study. NeuroImage: Clinical. 2022;36:103253. doi:10.1016/j.nicl.2022.103253

44. DiCiccio TJ, Efron B. Bootstrap confidence intervals. Statist Sci. 1996;11(3). doi:10.1214/ss/1032280214

45. Lee MD, Wagenmakers EJ. Bayesian Cognitive Modeling: A Practical Course. 1st ed. Cambridge University Press; 2014. doi:10.1017/CBO9781139087759

46. Griffith JW, Sokol CL, Luster AD. Chemokines and Chemokine Receptors: Positioning Cells for Host Defense and Immunity. Annu Rev Immunol. 2014;32(1):659–702. doi:10.1146/annurev-immunol-032713-120145

47. Turner MD, Nedjai B, Hurst T, Pennington DJ. Cytokines and chemokines: At the crossroads of cell signalling and inflammatory disease. Biochimica et Bio-physica Acta (BBA) - Molecular Cell Research. 2014;1843(11):2563–2582. doi:10.1016/j.bbamcr.2014.05.014

48. Momenilandi M, Lévy R, Sobrino S, et al. FLT3L governs the development of partially overlapping hematopoietic lineages in humans and mice. Cell. 2024;187(11):2817–2837.e31. doi:10.1016/j.cell.2024.04.009

49. Brkic M, Balusu S, Libert C, Vandenbroucke RE. Friends or Foes: Matrix Met-alloproteinases and Their Multifaceted Roles in Neurodegenerative Diseases. Vallières L, ed. Mediators of Inflammation. 2015;2015(1):620581. doi:10.1155/2015/620581

50. Cash DM, Bocchetta M, Thomas DL, et al. Patterns of gray matter atrophy in genetic frontotemporal dementia: results from the GENFI study. Neurobiology of Aging. 2018;62:191–196. doi:10.1016/j.neurobiolaging.2017.10.008

51. Whitwell JL, Weigand SD, Boeve BF, et al. Neuroimaging signatures of fronto-temporal dementia genetics: C9ORF72, tau, progranulin and sporadics. Brain. 2012;135(3):794–806. doi:10.1093/brain/aws001

52. Sogorb-Esteve A, Swift IJ, Woollacott IOC, Warren JD, Zetterberg H, Rohrer JD. Differential chemokine alteration in the variants of primary progressive aphasia—a role for neuroinflammation. J Neuroinflammation. 2021;18(1):224. doi:10.1186/s12974-021-02247-3

53. Loetscher M, Gerber B, Loetscher P, et al. Chemokine receptor specific for IP10 and mig: structure, function, and expression in activated T-lymphocytes. The Journal of experimental medicine. 1996;184(3):963–969. doi:10.1084/jem.184.3.963

54. Ellis SL, Gysbers V, Manders PM, et al. The Cell-Specific Induction of CXC Chemokine Ligand 9 Mediated by IFN-γ in Microglia of the Central Nervous System Is Determined by the Myeloid Transcription Factor PU.1. The Journal of Immunology. 2010;185(3):1864–1877. doi:10.4049/jimmunol.1000900

55. Zhou YQ, Liu DQ, Chen SP, et al. The Role of CXCR3 in Neurological diseases. CN. 2019;17(2):142–150. doi:10.2174/1570159X15666171109161140

56. Roos P, Von Essen MR, Nielsen TT, et al. Inflammatory markers of CHMP2B-mediated frontotemporal dementia. Journal of Neuroimmunology. 2018;324:136–142. doi:10.1016/j.jneuroim.2018.08.009

57. Singh S, Anshita D, Ravichandiran V. MCP-1: Function, regulation, and involvement in disease. International Immunopharmacology. 2021;101:107598. doi:10.1016/j.intimp.2021.107598

58. Yadav A, Saini V, Arora S. MCP-1: Chemoattractant with a role beyond immunity: A review. Clinica Chimica Acta. 2010;411(21-22):1570–1579. doi:10.1016/j.cca.2010.07.006

59. Lee WJ, Liao YC, Wang YF, Lin IF, Wang SJ, Fuh JL. Plasma MCP-1 and Cognitive Decline in Patients with Alzheimer’s Disease and Mild Cognitive Impairment: A Two-year Follow-up Study. Sci Rep. 2018;8(1):1280. doi:10.1038/s41598-018-19807-y

60. Broz P. Pyroptosis: molecular mechanisms and roles in disease. Cell Res. 2025;35(5):334–344. doi:10.1038/s41422-025-01107-6

61. Huang F, Zhu Y, Hsiao-Nakamoto J, et al. Longitudinal biomarkers in amyo-trophic lateral sclerosis. Ann Clin Transl Neurol. 2020;7(7):1103–1116. doi:10.1002/acn3.51078

62. Wijesinghe SS, Rowe JB, Mason HD, et al. Post-mortem validation of in vivo TSPO PET as a microglial biomarker. Brain. Published online February 26, 2025:awaf078. doi:10.1093/brain/awaf078

63. Vontobel DS, Lai KO, Bacioglu M, et al. TSPO PET binding in vivo reflects in-creased phagocytic microglia at post mortem in people with frontotemporal dementia. Pathology. Preprint posted online June 24, 2026. doi:10.64898/2026.06.22.26356176

64. Malpetti M, Rittman T, Jones PS, et al. In vivo PET imaging of neuroinflammation in familial frontotemporal dementia. J Neurol Neurosurg Psychiatry. 2021;92(3):319–322. doi:10.1136/jnnp-2020-323698

65. Bevan-Jones WR, Cope TE, Jones PS, et al. Neuroinflammation and protein aggregation co-localize across the frontotemporal dementia spectrum. Brain. 2020;143(3):1010–1026. doi:10.1093/brain/awaa033

66. Yilmaz A, Watson V, Else L, Gisslèn M. Cerebrospinal fluid maraviroc concentrations in HIV-1 infected patients. AIDS. 2009;23(18):2537–2540. doi:10.1097/QAD.0b013e328333ae0e

67. Rohrer JD, Nicholas JM, Cash DM, et al. Presymptomatic cognitive and neuroanatomical changes in genetic frontotemporal dementia in the Genetic Fronto-temporal dementia Initiative (GENFI) study: a cross-sectional analysis. The Lancet Neurology. 2015;14(3):253–262. doi:10.1016/S1474-4422(14)70324-2

68. Gaudet P, Michel PA, Zahn-Zabal M, et al. The neXtProt knowledgebase on human proteins: 2017 update. Nucleic Acids Res. 2017;45(D1):D177–D182. doi:10.1093/nar/gkw1062

69. Uhlén M, Fagerberg L, Hallström BM, et al. Tissue-based map of the human proteome. Science. 2015;347(6220):1260419. doi:10.1126/science.1260419

70. Aebersold R, Agar JN, Amster IJ, et al. How many human proteoforms are there? Nat Chem Biol. 2018;14(3):206–214. doi:10.1038/nchembio.2576

71. Bevan-Jones WR, Bevan-Jones WR, Cope TE, et al. In vivo evidence for pre-symptomatic neuroinflammation in a MAPT mutation carrier. Annals of clinical and translational neurology. 2019;6(2):373–378. doi:10.1002/acn3.683

