## Supplementary Material, Tables and Figures for "Atrophy-related inflammation CSF markers predict clinical progression in genetic frontotemporal dementia": CSF_GENFI_Manuscript_Supplementary_Material.docx

*Cambridge CB2 0SZ*

*United Kingdom*

**Supplementary methods**

**Clinical Outcomes**

To evaluate clinical decline, we considered the Sum of Boxes score from the CDR+NACC FTLD, as it has been widely used in FTD clinical trials as primary clinical outcome^1^ ^2^. The CDR® +NACC FTLD captures eight domains, including memory, orientation, judgement and problem-solving, community affairs, home and hobbies, personal care, overall behaviour, and overall language, which are particularly affected in FTD phenotypes^3^ ^4^ ^3–5^. The CDR® +NACC FTLD is administered in a semi-structured format involving both the person with FTD and the carer. A trained clinician assigns a score to each domain based on severity [using 0 (absent), 0.5 (questionable/very mild), 1 (mild), 2 (moderate), and 3 (severe)] ^6^. To compute the sum of boxes, all scores from the eight domains are summed, resulting in the CDR® +NACC-FTLD-SB (referred to as CDR-SB onwards). The CDR-SB ranges from 0-24, where higher scores indicate more severe symptomology ^6^.

Executive decline was indexed using the Wechsler Adult Intelligence Scale-Revised Digit Symbol Substitution Task (DSST)^7^, chosen for its strong test-retest reliability and sensitivity in presymptomatic stages. The DSST is a paper-and-pencil neuropsychological test that requires the participant to draw symbols corresponding to digits and match a pattern of symbols to numbers over a 90-second interval, resulting in continuous scores for correct pairings ranging from 100 (no executive dysfunction) to 0 (executive dysfunction).

**MRI processing**

MRI scans were bias-field corrected, and the whole brain was parcellated utilising the geodesic information flow (GIF) algorithm ^8^, and bilateral regions of interest (ROI) were computed; deeper cortical and limbic structures included the cingulate and the insula. Specialised FreeSurfer modules were employed to estimate the volumes of the cerebellum ^9^ and the brainstem ^10^. All segmentations underwent visual quality controls by the GENFI imaging core Team. Additionally, Total Intracranial Volume (TIV) ^11^ was calculated with SPM12 v6470 (Statistical Parametric Mapping, Wellcome Trust Centre for Neuroimaging, London, UK) under Matlab R2014b (Math Works, Natick, MA, USA). To account for head size, the 30 ROIs were divided by TIV, yielding ratios used in subsequent analysis.

**CSF sample collection and preprocessing**

CSF proteins were measured using the Olink® Target 96 platform, which employs multiplex panels based on Proximity Extension Assay (PEA) technology (Olink® Proteomics Inc., part of Thermo Fisher Scientific, Waltham, MA, USA). Further details on the PEA technology are described elsewhere^12^ ^13^. The concentrations of 92 proteins (inflammation panel) are reported as Olink’s relative protein quantification unit, Normalised Protein Expression (NPX), on a log2 scale. Information on assay characteristics and quality control procedures can be found on Olink’s website (<https://olink.com/>).

In the proteomics dataset, a small number of protein values (< 0.1 % of total measurements) were missing due to falling below the limit of detection (LOD). The below LOD values were imputed using the “impute.MinProb” (q=0.01) function from the imputeLCMD package ^14^ from the lower observed distribution. To examine whether batch effects were present across GENFI sites in the CSF data, we employed two complementary approaches. First, a PCA score plot (components 1 and 2) was visually inspected for systematic clustering by site using the “pca” function from the mixOmics package. Second, average silhouette width (ASW) ^15^ using the “silhouette” function from the cluster package. Silhouette coefficients were computed for each participant using Euclidean distances between normalised NPX values, with GENFI site as the grouping variable. The raw ASW score was transformed to a batch ASW score using 1 - |raw ASW| ^16^, yielding a metric ranging from 0 to 1, where values approaching 1 indicate the absence of batch effects. Prior to investigating the inflammation-brain covariance axis, we removed outliers to minimise the influence of extreme NPX values, which are most likely attributable to methodological or technical variations rather than true biological signal. For each protein column, we calculated the sample mean and standard deviation and flagged any participants with NPX values 5 SDs above or below the mean as outliers, a threshold used in previous proteomic studies ^17^. Participants with at least one flagged outlier value were excluded (n=11 (controls = 6, presymptomatic carriers = 3, symptomatic carriers = 2), 4.4 % of total sample), yielding a sample size of n=234. Here, we selected whole-participant exclusion as the more rigorous approach, rather than value-level replacement, because we deployed downstream multivariate statistical analyses (regularised Canonical Correlation Analysis (rCCA) and PCA) that rely on the covariance matrix ^18^. If flagged values were deleted and subsequently imputed, the resulting missingness would be directly determined by the magnitude of the value itself. This constitutes a missing-not-at-random mechanism, violating the missing-at-random assumption on which methods such as MICE depend ^19^. Simple substitution approaches (for example, mean or median imputation) would compress the variance-covariance structure ^20^. Both CCA and PCA are highly sensitive to such distortions ^21^, and their solutions could be compromised if derived from inappropriately imputed matrices. Whole-participant exclusion, therefore, represented the most statistically conservative and scientifically robust approach.

**Batch Effects**

PCA score plot derived from the CSF NPX values (components 1 and 2) was visually inspected for systematic clustering by site (Supplementary Figure 1), and a transformed ASW of 0.89. Both approaches converged, indicating no substantial batch effects across sites in the protein data.

**Regularised Canonical Correlation Analysis (rCCA)**

The objective of rCCA is to find the link between two multivariate matrices. In our context, we investigated the link between the CSF data set (X, 92 (proteins) x 234 (participants) matrix), regional brain values (Y, 30 (regional brain value) x 243 (participants) matrix), by assigning linear weights to matrix-specific features that map onto pairs of latent factors known as canonical variates (CV)^22^ (protein-CV [MRI weighted] and MRI-CV [protein weighted]) that are optimised to maximally correlate with each other. Prior to rCCA, we z-scored both X and Y matrices and applied an equal regularisation term ($\lambda=0.01$) to reduce the risk of overfitting. It is important to note that the interpretation of the modality-specific canonical variates derived by rCCA from proteomics and MRI data differs from other dimensionality reduction techniques, such as PCA. The rCCA algorithm is optimised to assign weights to each modality-specific canonical variate that maximise the correlation between the two modalities, rather than to reduce the dimensionality of the data in each modality independently. Hence, yielding participant scores (protein-CV and MRI-CV) with shared variance across proteomics and MRI data.

**Residualisation**

To isolate the protein-only signal from the atrophy-linked protein profiles (protein-CV), we residualised the 92 proteins by fitting 92 linear models (original protein~protein-CV) and then extracted the residuals. The residuals were normalised, and a PCA using the “prcomp” function (stats package) was applied to reduce 92 protein variables into principal components (PCs). First, the PCA analysis was performed including data from the symptomatic carrier group only as the previous analyses showed that protein-CV was statistically significantly higher than presymptomatic carriers and controls, and presymptomatic carriers were not statistically different from controls. To ensure PCA was appropriate, we applied Bartlett’s test for sphericity to assess whether the multidimensional data could be meaningfully compressed ^17^ using the “bart_spher” function from the REDaS package applied to the correlation matrix. The inclusion criteria for the number of retained PC for 2^nd^ level analysis were (a) components with eigenvalues > 1 (Kaiser criterion); (b) explained variance of >10%. Then, component scores were included in linear models to investigate the association between isolated protein signals and clinical decline and executive functioning. Next, to contextualise the residual structure, a separate PCA was performed on the normalised CSF NPX protein values for the presymptomatic carrier and control groups. The loading structures from the retained PCs (PCA on residualised within symptomatic carriers and a PCA on unresidualised in presymptomatic carriers/controls) were compared using Pearson's correlation.

**Supplementary Results**

**Result 1.1**

To further investigate prodromal changes, we also examined neurofilament light chain (NfL) abundance, a proxy marker for neurodegeneration, which is statistically significantly increased in prodromal (p = 0.03), and in phenoconverted stages (p<0.001, Main text Figure 3B). A similar pattern also emerges with CXCL9 (the highest loaded protein in the rCCA), showing statistically significant increases in prodromal (p=0.04) and phenoconverted (p<0.001) stages (Main text Figure 3E).

**Result 1.2**

The omnibus interaction term between CXCL9 and the re-stratified CDR groups in the parametric model was statistically significant (F(3,190)=5.07, p=0.002), associated with CDR slope, which is in concordance with the bootstrapped model, where the interaction terms were statistically significant in phenoconverted participants with worse clinical progression (β=0.29, CI:[0.07,0.52, Main text Figure 4F]) contrary to the remaining groups which was not associated with CDR slopes (controls β=-0.08, CI:[-0.19,0.06], Asymptomatic β=0.03, CI:[-0.13,0.17], Prodromal β=0.09, CI:[-0.15,0.29]).

**Result 1.3**

The omnibus interaction term between NfL levels and the re-stratified CDR groups in the parametric model was statistically significant (F(3,170)=13.72, p<0.001), associated with CDR slope, which is in concordance with the bootstrapped model, where the interaction terms were statistically significant, interestingly, for NfL, both asymptomatic (β=0.35, CI:[0.04,0.84]) and phenoconverted (β=1.31, CI:[0.93,1.73]) participants were statistically significantly associated with CDR slopes whereas the remaining two groups were not (Controls β=-0.09, CI:[-0.38,0.14], Prodromal β=0.87, CI:[-0.06,1.80] Main text Figure 4B).

**Result 1.4**

The omnibus interaction term between CXCL9 levels and the re-stratified CDR groups in the parametric model was not statistically significant (F(3,189)=0.48, p=0.69), associated with DSST scores, which is in concordance with the bootstrapped model, where the interaction terms were also not statistically significant in phenoconverted participants (β=-1.55, CI:[-8.48,4.40] Main text Figure 4H), whereas this was not the case for the remaining groups (controls β=0.33, CI:[-4.18,4.34], Asymptomatic β=1.44, CI:[-3.88,6.12], Prodromal β=2.81, CI:[-8.37,13.17]).

**Result 1.5**

The omnibus interaction term between NfL levels and the re-stratified CDR groups in the parametric model was statistically significant (F(3,166)=5.44, p=0.001), associated with DSST scores, which is in concordance with the bootstrapped model, where the interaction terms were statistically significant in phenoconverted participants associations between NfL levels and DSST scores (β=-33.63, CI:[-50.83,-15.50] Main text Figure 4D), whereas this was not the case for the remaining groups (controls β=12.20, CI:[-5.24,27.99], Asymptomatic β=-11.30, CI:[-31.96,10.04], Prodromal β=-23.77, CI:[-45.54,13.69]).

**Supplementary Figures**

Supplementary Figure 1: Principal Component Analysis (PCA) of protein values to evaluate potential batch effects. Individual data points are projected onto the first two principal components (PC1, 37% explained variance; PC2, 9% explained variance), with marginal plots displaying the overall density distributions. Samples are colour-coded by GENFI sites and shaped by disease status (HC = healthy controls, PMC = presymptomatic mutation carriers, SMC = symptomatic mutation carriers). The high degree of overlap among distributions and the lack of discrete, site-specific clustering strongly indicate the absence of systematic batch effects.

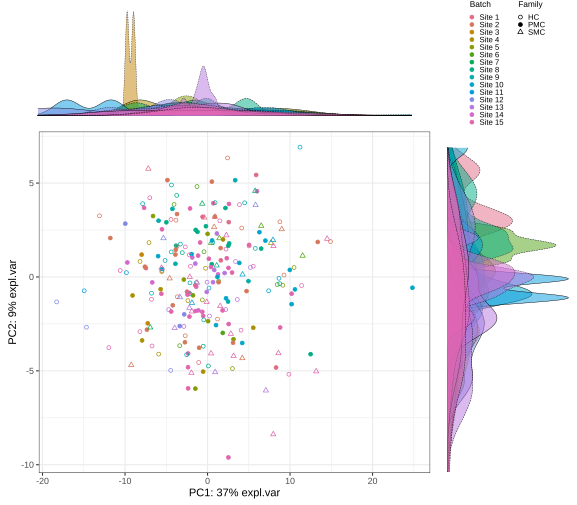

Supplementary Figure 2: Cross-sectional differences and longitudinal trajectories in CDR plus NACC FTLD-SB and DSST across the diagnostic groups. (A) Left: Violin plots of CDR plus NACC FTLD Sum of Boxes (CDR+NACC-FTLD-SB) by disease status (HC, healthy non-carriers; PMC, presymptomatic mutation carriers; SMC, symptomatic mutation carriers. Points represent group means. SMC is significantly elevated over both (p < 0.0001). Right: Individual longitudinal trajectories of CDR+NACC-FTLD-SB, coloured by group (blue = HC, green = PMC, orange = SMC). (B) Left: Violin plots of Digit Symbol Substitution Test (DSST) by disease status (HC = healthy non-carriers; PMC = presymptomatic mutation carriers; SMC = symptomatic mutation carriers). Points represent group means. SMC is significantly lower over both (p < 0.0001). Right: Individual longitudinal trajectories of DSST, coloured by group (blue = HC, green = PMC, orange = SMC).

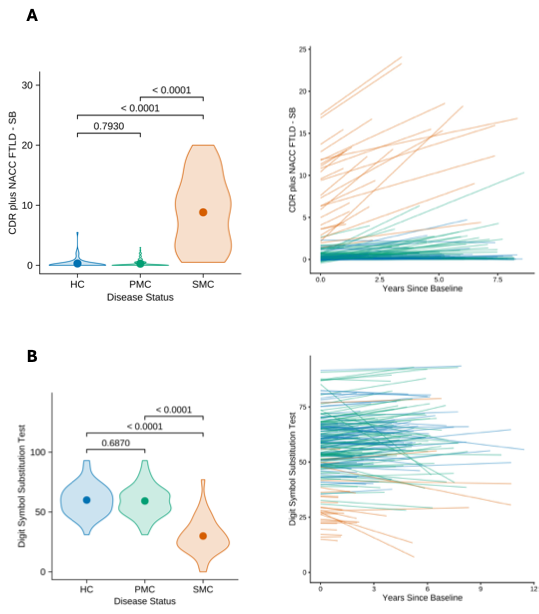

Supplementary Figure 3. Comparative analysis of principal component loading profiles of inflammation proteins with and without residualisation. Heatmap contrasting the individual protein loadings of the first principal component (PC1) derived from unresidualised protein values in the presymptomatic mutation carrier and healthy control groups (PC1 - unresidualised in PMC/HC, upper) against the PC1 loadings derived from residualised values within the symptomatic mutation carrier group (PC1 - residualised within SMC, lower). Individual proteins are displayed along the x-axis, with colour intensity corresponding to loading magnitude.

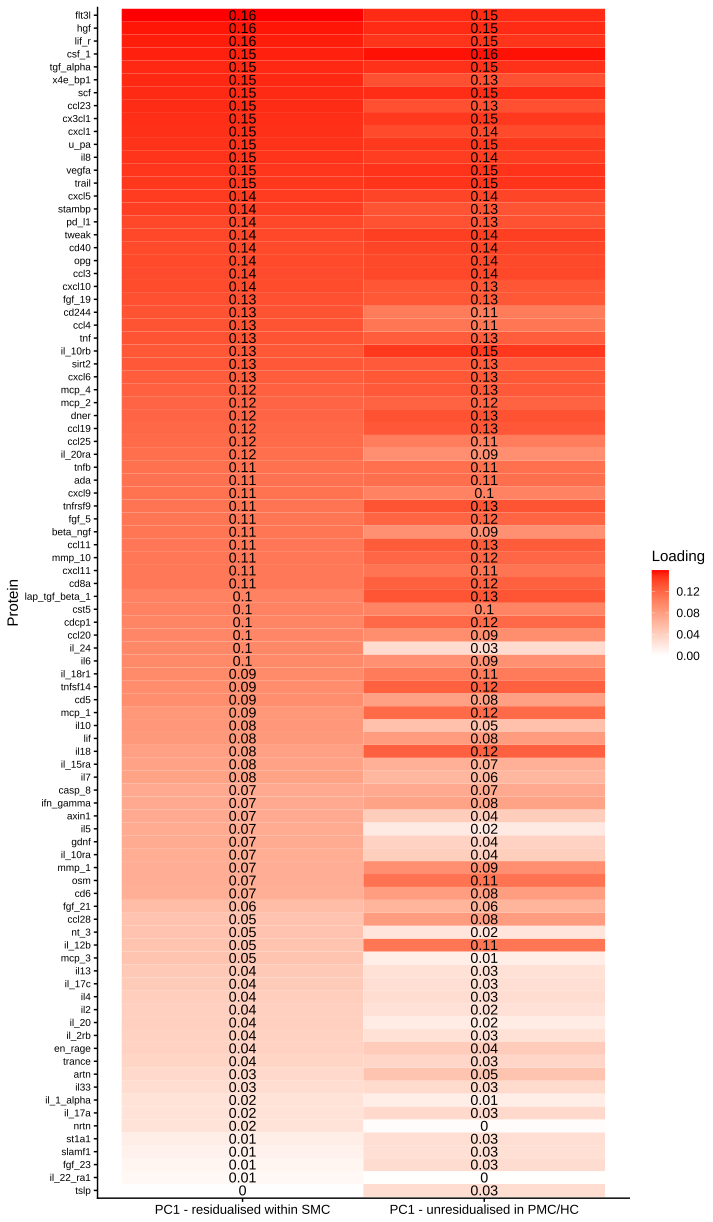

Supplementary Figure 4. Comparative analysis of principal component loading profiles and scores between disease groups. (A) Scatter plot demonstrating a strong, positive correlation between the PC1 protein loading coefficients derived from the unresidualised PMC/HC cohort (y-axis) and those from the residualised SMC cohort (x-axis). The solid black line represents the line of best fit, with the shaded area indicating the 95% confidence interval, illustrating the incredibly high structural preservation of the inflammatory protein profile across the cohort. (B) Boxplot comparing the distribution of PC1 scores between healthy controls (HC, teal-green) and presymptomatic mutation carriers (PMC, orange).

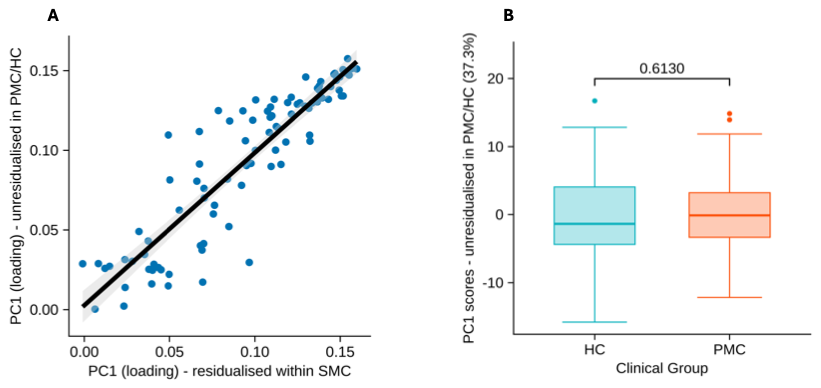

Supplementary Figure 5. Multimodal biomarker profiles across genetic groups along the re-stratified clinical continuum. Mutation carriers were re-stratified by baseline CDR plus NACC FTLD-SB (Asymptomatic: CDR-SB = 0; Prodromal: CDR-SB = 0.5; Phenoconverted: CDR-SB ≥ 1) and further stratified by mutation group (C9orf72, GRN, MAPT); healthy controls are not shown. Individual datapoints are not plotted. Violin plots represent data density distributions across the clinical continuum; black dots track group-wise means. Within each biomarker (row), the y-axis is held constant across clinical stages to permit visual comparison of stage-related change. Group sizes were n = 35/29/14 (C9orf72/GRN/MAPT) at the Asymptomatic, 9/4/6 at the Prodromal, and 21/14/9 at the Phenoconverted stage for Protein CV, MRI CV and CXCL9; and n = 27/26/12, 9/3/5 and 18/14/7, respectively, for log10 NfL.

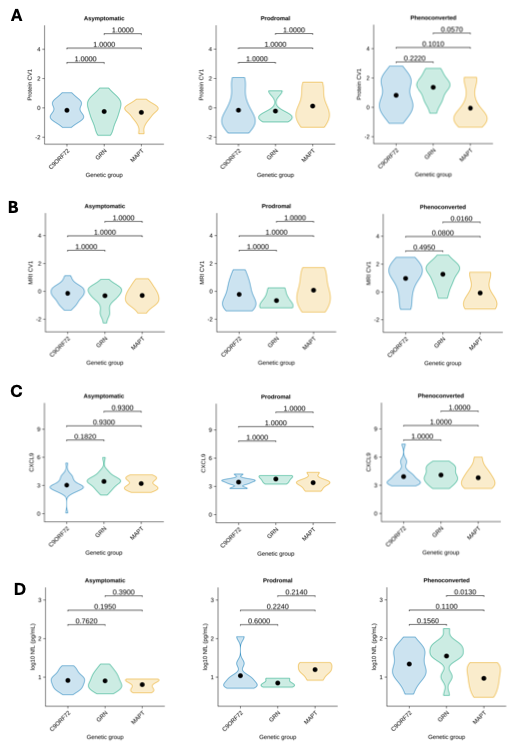

**Supplementary Tables**

Supplementary Table 1. Loadings of protein variables (red) and regional MRI

values (blue) on the first rCCA component.

| **Category** | **Variable** | **Loading (contribution to CCA Component)** |
| --- | --- | --- |
| Protein | cxcl9 | 0.562 |
| Protein | flt3l | 0.480 |
| Protein | ccl3 | 0.451 |
| Protein | mcp_1 | 0.404 |
| Protein | lap_tgf_beta_1 | 0.385 |
| Protein | ccl25 | 0.376 |
| Protein | mmp_10 | 0.366 |
| Protein | opg | 0.346 |
| Protein | sirt2 | 0.308 |
| Protein | il18 | 0.306 |
| Protein | il_15ra | 0.291 |
| Protein | il7 | 0.275 |
| Protein | cd244 | 0.264 |
| Protein | stambp | 0.260 |
| Protein | ccl28 | 0.256 |
| Protein | il8 | 0.248 |
| Protein | il_10rb | 0.247 |
| Protein | cd40 | 0.239 |
| Protein | osm | 0.237 |
| Protein | tnfrsf9 | 0.235 |
| Protein | x4e_bp1 | 0.220 |
| Protein | hgf | 0.212 |
| Protein | cxcl1 | 0.208 |
| Protein | pd_l1 | 0.193 |
| Protein | scf | 0.189 |
| Protein | mcp_4 | 0.184 |
| Protein | fgf_21 | 0.179 |
| Protein | ccl23 | 0.172 |
| Protein | cxcl10 | 0.167 |
| Protein | il_20 | 0.166 |
| Protein | slamf1 | 0.157 |
| Protein | ccl11 | 0.154 |
| Protein | cst5 | 0.153 |
| Protein | csf_1 | 0.145 |
| Protein | axin1 | 0.145 |
| Protein | tnf | 0.141 |
| Protein | cxcl6 | 0.140 |
| Protein | cxcl5 | 0.138 |
| Protein | il_10ra | 0.132 |
| Protein | tnfb | 0.132 |
| Protein | il_18r1 | 0.124 |
| Protein | mcp_2 | 0.117 |
| Protein | il13 | 0.109 |
| Protein | il10 | 0.105 |
| Protein | il5 | 0.103 |
| Protein | casp_8 | 0.098 |
| Protein | ccl4 | 0.095 |
| Protein | cxcl11 | 0.095 |
| Protein | st1a1 | 0.087 |
| Protein | il6 | 0.079 |
| Protein | ifn_gamma | 0.078 |
| Protein | tnfsf14 | 0.077 |
| Protein | dner | 0.077 |
| Protein | tslp | 0.075 |
| Protein | lif | 0.074 |
| Protein | il_2rb | 0.074 |
| Protein | ccl20 | 0.072 |
| Protein | cdcp1 | 0.069 |
| Protein | il_22_ra1 | 0.066 |
| Protein | trance | 0.065 |
| Protein | lif_r | 0.053 |
| Protein | ada | 0.039 |
| Protein | beta_ngf | 0.037 |
| Protein | mcp_3 | 0.037 |
| Protein | tweak | 0.036 |
| Protein | cx3cl1 | 0.033 |
| Protein | tgf_alpha | 0.032 |
| Protein | il_17c | 0.027 |
| Protein | nt_3 | 0.018 |
| Protein | trail | 0.017 |
| Protein | u_pa | 0.015 |
| Protein | ccl19 | 0.008 |
| Protein | il33 | 0.005 |
| Protein | cd8a | 0.001 |
| Protein | il_12b | -0.002 |
| Protein | en_rage | -0.002 |
| Protein | il_17a | -0.002 |
| Protein | il_1_alpha | -0.003 |
| Protein | cd6 | -0.004 |
| Protein | gdnf | -0.014 |
| Protein | nrtn | -0.015 |
| Protein | il2 | -0.024 |
| Protein | il4 | -0.045 |
| Protein | cd5 | -0.045 |
| Protein | fgf_19 | -0.053 |
| Protein | vegfa | -0.055 |
| Protein | fgf_23 | -0.064 |
| Protein | il_24 | -0.074 |
| Protein | il_20ra | -0.089 |
| Protein | mmp_1 | -0.103 |
| Protein | artn | -0.107 |
| Protein | fgf_5 | -0.207 |
| MRI | right_insula_volume | -0.848 |
| MRI | left_insula_volume | -0.836 |
| MRI | lateral_temp_right | -0.828 |
| MRI | lateral_temp_left | -0.819 |
| MRI | dlpfc_left | -0.748 |
| MRI | lateral_parietal_right | -0.732 |
| MRI | vmpfc_left | -0.705 |
| MRI | motor_right | -0.693 |
| MRI | orbitofrontal_left | -0.673 |
| MRI | motor_left | -0.662 |
| MRI | lateral_parietal_left | -0.661 |
| MRI | vmpfc_right | -0.659 |
| MRI | supra_temp_right | -0.656 |
| MRI | dlpfc_right | -0.650 |
| MRI | left_cingulate_volume | -0.627 |
| MRI | medial_parietal_left | -0.620 |
| MRI | medial_parietal_right | -0.619 |
| MRI | supra_temp_left | -0.580 |
| MRI | orbitofrontal_right | -0.557 |
| MRI | right_cingulate_volume | -0.550 |
| MRI | sensory_right | -0.542 |
| MRI | sensory_left | -0.533 |
| MRI | left_occipital_lobe_volume | -0.524 |
| MRI | right_occipital_lobe_volume | -0.505 |
| MRI | medial_temp_left | -0.477 |
| MRI | total_cerebellum | -0.462 |
| MRI | temporal_pole_right | -0.457 |
| MRI | medial_temp_right | -0.420 |
| MRI | temporal_pole_left | -0.366 |
| MRI | brain_stem | -0.119 |

Supplementary Table 2. Bootstrapped linear regression models mapping baseline protein CV scores against clinical and cognitive outcomes in presymptomatic mutation carriers and healthy controls. Regression coefficients and 95% bias-corrected and accelerated confidence intervals (CI 95% BCa) are reported individually for the presymptomatic mutation carrier (PMC) and healthy control (HC) groups. Across all cross-sectional and longitudinal (slope) clinical and cognitive outcomes.

| **Outcome** | **Group** | **β** | **95% CI** |
| --- | --- | --- | --- |
| CDR | PMC | -0.07 | [-0.26, 0.07] |
| CDR Slope | PMC | 0.04 | [-0.01, 0.17] |
| DSST | PMC | 0.91 | [-2.99, 4.59] |
| DSST Slope | PMC | -0.20 | [-0.53, 0.08] |
| CDR | HC | -0.14 | [-0.55, 0.08] |
| CDR Slope | HC | 0.01 | [-0.04, 0.07] |
| DSST | HC | 2.65 | [-2.65, 7.65] |
| DSST Slope | HC | -0.22 | [-0.53, 0.08] |

References:

1.	Assogna M, Di Lorenzo F, Bonnì S, et al. Phase 2 study of palmitoylethanolamide combined with luteoline in frontotemporal dementia patients. *Brain Communications*. 2025;7(2):fcaf080. doi:10.1093/braincomms/fcaf080

2.	Koch G, Assogna M, Gadola Y, et al. Safety and efficacy of rotigotine in patients with frontotemporal dementia: a phase 2, double-blind, randomized, placebo-controlled, multicenter trial. *The Lancet Regional Health - Europe*. 2025;57:101409. doi:10.1016/j.lanepe.2025.101409

3.	Morris JC. The Clinical Dementia Rating (CDR): Current version and scoring rules. *Neurology*. 1993;43(11):2412. doi:10.1212/WNL.43.11.2412-a

4.	Miyagawa T, Brushaber D, Syrjanen J, et al. Use of the CDR® plus NACC FTLD in mild FTLD: Data from the ARTFL/LEFFTDS consortium. *Alzheimer’s &amp; Dementia*. 2020;16(1):79-90. doi:10.1016/j.jalz.2019.05.013

5.	Knopman DS, Weintraub S, Pankratz VS. Language and behavior domains enhance the value of the clinical dementia rating scale. *Alzheimer’s &amp; Dementia*. 2011;7(3):293-299. doi:10.1016/j.jalz.2010.12.006

6.	Peakman G, Russell LL, Convery RS, et al. Comparison of clinical rating scales in genetic frontotemporal dementia within the GENFI cohort. *J Neurol Neurosurg Psychiatry*. 2022;93(2):158-168. doi:10.1136/jnnp-2021-326868

7.	Wechsler D. WAIS-R: Manual: Wechsler adult intelligence scale-revised. Published online 1981.

8.	Cardoso MJ, Modat M, Wolz R, et al. Geodesic Information Flows: Spatially-Variant Graphs and Their Application to Segmentation and Fusion. *IEEE Trans Med Imaging*. 2015;34(9):1976-1988. doi:10.1109/TMI.2015.2418298

9.	Diedrichsen J, Balsters JH, Flavell J, Cussans E, Ramnani N. A probabilistic MR atlas of the human cerebellum. *NeuroImage*. 2009;46(1):39-46. doi:10.1016/j.neuroimage.2009.01.045

10.	Iglesias JE, Van Leemput K, Bhatt P, et al. Bayesian segmentation of brainstem structures in MRI. *NeuroImage*. 2015;113:184-195. doi:10.1016/j.neuroimage.2015.02.065

11.	Malone IB, Leung KK, Clegg S, et al. Accurate automatic estimation of total intracranial volume: A nuisance variable with less nuisance. *NeuroImage*. 2015;104:366-372. doi:10.1016/j.neuroimage.2014.09.034

12.	Del Campo M, Peeters CFW, Johnson ECB, et al. CSF proteome profiling across the Alzheimer’s disease spectrum reflects the multifactorial nature of the disease and identifies specific biomarker panels. *Nat Aging*. 2022;2(11):1040-1053. doi:10.1038/s43587-022-00300-1

13.	van der Ende EL, In ’t Veld SGJG, Hanskamp I, et al. CSF proteomics in autosomal dominant Alzheimer’s disease highlights parallels with sporadic disease. *Brain*. 2023;146(11):4495-4507. doi:10.1093/brain/awad213

14.	Lazar C, Gatto L, Ferro M, Bruley C, Burger T. Accounting for the Multiple Natures of Missing Values in Label-Free Quantitative Proteomics Data Sets to Compare Imputation Strategies. *J Proteome Res*. 2016;15(4):1116-1125. doi:10.1021/acs.jproteome.5b00981

15.	Rousseeuw PJ. Silhouettes: A graphical aid to the interpretation and validation of cluster analysis. *Journal of Computational and Applied Mathematics*. 1987;20:53-65. doi:10.1016/0377-0427(87)90125-7

16.	Tran HTN, Ang KS, Chevrier M, et al. A benchmark of batch-effect correction methods for single-cell RNA sequencing data. *Genome Biol*. 2020;21(1):12. doi:10.1186/s13059-019-1850-9

17.	Malpetti M, Swann P, Tsvetanov KA, et al. Blood inflammation relates to neuroinflammation and survival in frontotemporal lobar degeneration. *Brain*. 2025;148(2):493-505. doi:10.1093/brain/awae269

18.	Zhuang X, Yang Z, Cordes D. A technical review of canonical correlation analysis for neuroscience applications. *Human Brain Mapping*. 2020;41(13):3807-3833. doi:10.1002/hbm.25090

19.	Van Buuren S. *Flexible Imputation of Missing Data, Second Edition*. 2nd ed. Chapman and Hall/CRC; 2018. doi:10.1201/9780429492259

20.	Little R, Rubin D. *Statistical Analysis with Missing Data, Third Edition*. 1st ed. Wiley; 2019. doi:10.1002/9781119482260

21.	Luo L, Wang W, Bao S, Peng X, Peng Y. Robust and sparse canonical correlation analysis for fault detection and diagnosis using training data with outliers. *Expert Systems with Applications*. 2024;236:121434. doi:10.1016/j.eswa.2023.121434

22.	Wang HT, Smallwood J, Mourao-Miranda J, et al. Finding the needle in a high-dimensional haystack: Canonical correlation analysis for neuroscientists. *NeuroImage*. 2020;216:116745. doi:10.1016/j.neuroimage.2020.116745

23.	Bocchetta M, Todd EG, Peakman G, et al. Differential early subcortical involvement in genetic FTD within the GENFI cohort. *NeuroImage: Clinical*. 2021;30:102646. doi:10.1016/j.nicl.2021.102646

24.	Rohrer JD, Nicholas JM, Cash DM, et al. Presymptomatic cognitive and neuroanatomical changes in genetic frontotemporal dementia in the Genetic Frontotemporal dementia Initiative (GENFI) study: a cross-sectional analysis. *The Lancet Neurology*. 2015;14(3):253-262. doi:10.1016/S1474-4422(14)70324-2

25.	Miyagawa T, Brushaber D, Syrjanen J, et al. Utility of the global CDR® plus NACC FTLD rating and development of scoring rules: Data from the ARTFL/LEFFTDS Consortium. *Alzheimer’s &amp; Dementia*. 2020;16(1):106-117. doi:10.1002/alz.12033
